# Diversity of antibiotic resistance genes increases in urbanized lakes: a multi-tool screening

**DOI:** 10.64898/2026.01.25.26344777

**Authors:** Pau De Yebra, Luca Zoccarato, John A Galindo, Daniela Numberger, Nafi’u Abdulkadir, Hans-Peter Grossart, Alex D. Greenwood

**Affiliations:** Leibniz Institute for Zoo and Wildlife Research, Alfred-Kowalke-Strasse 17, 10315 Berlin, Germany; Leibniz Institute of Freshwater Ecology and Inland Fisheries, Dept, of Plankton and Microbial Ecology, Zur alten Fischerhütte 2, 16775 Stechlin, Germany; University of Potsdam, Institute of Biochemistry and Biology, Karl-Liebknecht-Strasse 24-25, 14476 Potsdam, Germany; Institute of Computational Biology, University of Natural Resources and Life Sciences (BOKU), Muthgasse 18, Vienna 1190, Austria; Core Facility Bioinformatics, University of Natural Resources and Life Sciences (BOKU), Muthgasse 18, Vienna 1190, Austria; Sokoto State University, Department of Microbiology, Birnin Kebbi Rd, 852101, Sokoto, Nigeria; Berlin-Brandenburg Institute of Advanced Biodiversity Research (BBIB), Altensteinstrasse 32, 14195 Berlin, Germany; Freie Universität Berlin, Department of Veterinary Medicine, Institute for Virology, Robert von Ostertag-Strasse 7-13, 14163 Berlin, Germany

**Author notes:** **To whom correspondence should be addressed: Professor Alex D. Greenwood**, Leibniz Institute for Zoo and Wildlife Research, Department of Wildlife Diseases, Alfred-Kowalke-Str. 17, 10315 Berlin, Germany, **Professor Hans-Peter Grossart,** Leibniz Institute of Freshwater Ecology and Inland Fisheries, Deptartment of Plankton and Microbial Ecology, Zur alten Fischerhütte 2, 16775 Stechlin, Germany.

**Keywords:** Antimicrobial resistance (AMR), Antibiotic resistance genes (ARGs), Deep sequencing, Wastewater treatment plant (WWTP), Urban Lake

## Abstract

Antimicrobial resistance (AMR) is a growing global public health threat projected to cause up to 10 million deaths annually by 2050 if no immediate action is taken. While misuse and overuse of antibiotics are the main drivers of increasing AMR, the eco-evolutionary dynamics of AMR in the environment – particularly across the urban-rural continuum – remain poorly understood. Using shotgun sequencing, we investigated urban, farm, and rural water sources in the Berlin-Brandenburg region to explore the distinctness or overlap of their antibiotic resistance gene (ARG) profiles and the potential impact of wastewater treatment plants (WWTP). ARGs were identified using multiple databases and five bioinformatic tools, combining sequence-based alignment and deep learning approaches. This multi-tool approach allowed for the detection of up to 18 AMR classes—more than any single tool alone. The multi-tool screening approach for ARGs, combined with the ABRicate algorithm, was superior to all single ARG tools and databases, detecting more AMR classes, allowing for biocide and metal resistance detection, while less sensitive for detection of aminocoumarin resistance genes. ARG diversity was higher in urban lake sediments, urban waters, and wastewater compared to rural lake sediments and water. Among all environments, urban lake water showed the highest overall ARG abundance, second only to wastewater, and this pattern held across all AMR classes, except for aminoglycoside resistance, which was most prevalent in rural lake sediments. The WWTP was unable to remove the circulating pool of ARGs, despite a decrease in unique ARGs in the outflow.

## 1. Introduction

Despite increasing information on microbial communities in urban water environments (Fan et al., 2022; Li et al., 2023; Numberger et al., 2022; Bai Y et al., 2022), it is still uncertain whether similar patterns govern the abundance and diversity of antimicrobial resistance genes (ARGs) (Kümmerer, 2004), as the impact of human activities can differ within the same biome (McInnes et al., 2021; Zhang et al., 2022b; Zhu et al., 2022). Urban water systems, such as lakes or rivers, have been described as reservoirs and major sources of ARGs from domestic, industrial, and hospital wastewater effluents (Quillaguamán et al., 2021; Jiang et al., 2022, 2024; Kraemer et al., 2022; Sabar et al., 2023). Urban lakes with high anthropogenic activity and industrial effluent exposure exhibit high concentrations of ARGs and heavy metals (Rajasekar et al., 2022). Rural environments instead show diverse ARG profiles, linked to land use, agricultural practices, and environmental factors (Du et al., 2025; Zeng et al., 2025). Urban areas often have higher overall ARG loads, but rural settings can harbor unique ARG profiles and high abundances in specific locations, such as agricultural areas or areas with proximity to animal husbandry or aquaculture (Manyi-Loh et al., 2018). While urban and rural areas exhibit different patterns and drivers, they can be interconnected through various pathways, including wastewater discharge, animal movement, and human activity.

Understanding ARG profiles and distribution in aquatic environments is crucial because of the major threat that antimicrobial resistance (AMR) poses to environmental, animal, and human health (UNEP, 2023). Recent evidence suggests that anthropogenic impact strongly influences the development, transmission, spread, and persistence of ARGs (Rzymski et al., 2024). AMR dynamics, particularly in urban aquatic environments exposed to activities such as wastewater treatment, aquaculture, clinical waste, or landfills, remain complex and poorly understood (UNEP, 2023). This is particularly true for WWTPs, which can act as hotspots for ARGs in the aquatic environment (Duarte et al., 2023; Honda et al., 2023).

Advances in sequencing technologies and bioinformatics have improved ARG detection and classification in clinical and environmental samples, including detection of mobile genetic elements (MGEs) such as plasmids, transposons, and proviruses integrated in host genomes, which can play a key role in AMR transmission. Tools like geNomad (Camargo et al., 2023) enable MGE prediction from sequencing data, while a range of ARG detection tools—based on sequence alignment (e.g., ABRicate, AMRFinderPlus, RGI, Staramr) or deep learning (e.g., DeepARG)—can be applied to short-read data (Marini et al., 2022; Wu et al., 2023).

Several studies have compared the performance and accuracy between different ARG tools and databases showing that they may not be suitable for detecting ARGs at low abundance (<1% of bacterial populations) (Wissel et al., 2023; Cooper et al., 2024). Different tools vary in detecting AMR classes due to differences in methodological approaches, databases, nomenclature, or their capacity to detect point mutations. Many databases are biased toward clinical resistance genes or clinical isolates (Papp and Solymosi, 2022; Xu et al., 2024), introducing bias when annotating environmental ARGs. For instance, the CARD database (Alcock et al., 2023) includes a higher diversity of microbial genera and AMR variants when compared to other databases, which could make the database better suited for environmental screening of ARGs.

Recent studies suggest that each method be better suited for detecting specific classes of ARGs (Marini et al., 2022). To this end, hAMRoaster has been developed to assess the performance of different ARG tools using a synthetic dataset from strains isolated from human tissues (Wissel et al., 2023). In a high resistance mock community test, all tools exhibited high precision and accuracy, but they still reported differences in ARG detection across different ARG drug classes. The main limitation is that similar comparisons of ARG screening tools for application to environmental samples are totally lacking (Goodarzi et al., 2022) and predicting environmental ARGs relying on a single ARG screening tool may miss some key AMR classes.

To better address the potential connectivity across urban and rural freshwater ecosystems and the knowledge gaps existing in characterizing ARG abundance and diversity, we performed shotgun metagenomic sequencing on water and sediment samples collected in the German states of Berlin and Brandenburg. We adopted an ensemble approach, combining 5 ARG screening tools and a total of 9 ARG tool-database, to overcome the bioinformatic limitations described above. We hypothesized that: (1) a multi-tool screening approach would detect a broader range of AMR classes than any single tool; (2) ARG abundance and diversity would be higher in urban lake water and sediment samples compared to rural ones; and (3) ARG abundance and diversity would differ significantly among WWTP inflow, outflow, and lake environments; (4) Unique profiles and diversity in ARGs characteristic of the different environments compared are linked to selective exposures; and (5) WWTP might not efficiently remove ARGs from circulating water or display a systematic bias.

## 2. Material and Methods

### 2.1 Sample collection

Details on the collection and grouping of environmental samples are provided in Section 1.1. of the Supplementary Material.

### 2.2 Illumina short-read sequencing

Details on the procedures and characteristics of Illumina short-read sequencing are provided in Section 1.2. of the Supplementary Material.

### 2.3. Bioinformatic analyses

The paired reads sequences from the Illumina platform were trimmed, filtered and PhiX and human sequences removed using BBtools v38.96 (Bushnell B. – sourceforge.net/projects/bbmap/). The resulting high-quality retained reads had a quality score >20. Reads were assembled into contigs using Megahit *v1*.*2*.*9 (parameters: --min-count 2, --k-list 21,41,61,81,99, --min-contig-len 200*) (Liu et al., 2014). Open reading frames (ORFs) were predicted from the metagenome assemblies using Prodigal (Hyatt et al., 2010) and a non-redundant ORF set was generated by merging all predicted ORFs and dereplicating using the SeqKit v2.8.2. A first clustering step was conducted by clustering the non-redundant ORF set with MMseqs2 v13.45111 (Steinegger and Söding, 2017) with the following parameters: –min-seq-id 0.95, -coverage 0.95, --cov-mode 1 and --cluster-mode 2. The sequences extracted were then pooled and a second round of clustering was executed to generate the final ORF catalog. Next, the high-quality reads from each sample were mapped to the ORF catalog using bowtie2 v2.4.5 (Langmead and Salzberg, 2012) and Samtools coverage v1.14 (Li et al., 2009) to compute the number of reads mapped.

A matrix based on the normalization of counts using RPKM-SCMG (Reads Per Kilobase of transcript per Million reads mapped and single copy marker gene) was computed by dividing the number of mapped reads by the ORF length, by 10^6^ and by the median count value of ORFs annotated as 10 universal single-copy phylogenetic marker genes (K06942, K01889, K01887, K01875, K01883, K01869, K01873, K01409, K03106, and K03110) (Salazar et al., 2019). The KEGG Orthology annotation was performed using KofamScan v1.3.0 (Aramaki et al., 2020). Therefore, the normalized ORF counts represent the average number of copies per cell.

### 2.4. Functional annotation of genes

The ORF catalog was annotated with eggNOG mapper v2.1.5 (Cantalapiedra et al., 2021) to retrieve the Clusters of Orthologous Groups of proteins (COGs). The rarefaction curve of the COGs (Fig. S2) indicates that the ORF catalog covers most of the COG diversity for the three different environments (water, sediment, and WWTP).

### 2.5. Antibiotic resistance gene (ARG) screening using multiple tools

The screening for antibiotic resistance genes (ARGs) and the following analyses were conducted over the ORF catalog. In order to achieve a robust and consistent ARG annotation, we conducted a multi-tool ARG screening using 5 ARG screening tools and a total of 9 combinations of the ARG tool databases with two different methodological approaches: Sequence alignment [AMRFinderplus-AMRFinderplusdb (Feldgarden et al., 2021), ABRicate (https://github.com/tseemann/ABRicate) with multiple databases, i.e. NCBI, MEGARes, ARG-ANNOT, CARD and ResFinder), RGI-CARD (McArthur et al., 2013) and staramr-ResFinder (Bharat et al., 2022)] and Deep learning [DeepARG-DeepARGDB (Arango-Argoty et al., 2018)]. More information on each specific tool and database is provided in Section 1.3. of the Supplementary Material.

Potential ARGs detected with sequence alignment and deep learning tools were included in the analysis if their identity and coverage values when aligned to the query sequence were higher than 80% (Arango-Argoty et al., 2018; Gupta et al., 2020; Abdulkadir et al., 2024; Cooper et al., 2024). In this manner, matches with low similarity were discarded from the analysis while ARG sequence variability was considered to allow for detection of ARG variants in the environment that may have diverged from the collection of clinical ARGs in databases.

Based on the number of ARG hits annotated by just one tool or more than one tool (Fig. S3), only those genes annotated by at least two out of the five AMR tools (DeepARG, AMRFinderplus, RGI, Staramr and ABRICATE considering all databases) were considered ARGs and were included in further analysis. Finally, the annotation was standardized based on the CARD nomenclature (Alcock et al., 2023). If two tools reported different AMR classes for a given ORF, the resulting annotation was defined as “ambiguous”. However, if one of the annotations belonged to “unclassified” or “multidrug” classes, while the other annotation/s agreed on a specific AMR class, then the precise annotation was assigned (see Section 1.4. of the Supplementary Material).

### 2.6. Taxonomic annotation of genes

For the taxonomic classification of all ARGs in the gene catalog we used Kraken version 2.1.3 Copyright 2013-2023 (https://ccb.jhu.edu/software/kraken2/) with the core_nt Database (https://benlangmead.github.io/aws-indexes/k2). Only classified “C” sequences were used for further analysis with the aim of linking the predicted taxa with the ARGs at the ORF level.

### 2.7. MGE sequence annotation

For the annotation of mobile genetic elements, i.e. plasmid and viral sequences, from the ORF non-redundant dataset, we used geNomad v1.8.1 (Camargo et al., 2023) with the following parameters: genomad end-to-end --cleanup --splits 8 genomad_db/ --threads 32 --verbose.

## 3. Results

### 3.1. AMR class detection

Up to 18 AMR classes were detected by the multi-tool approach (Fig. 1A) with the cutoff being ARGs detected by at least two tools. Diversity of AMR classes were found in descending order from wastewater treatment plant (WWTP) inflow (all 18 classes) > WWTP outflow (16 classes) > Müggel water (10 classes) > Weißer sediment (9 classes) > Dagow sediment (6 classes) = Stechlin sediment (6 classes) = Haus sediment (6 classes) = farm pond (6 classes) > Müggel sediment (4 classes) > Weißer water (2 classes), and no ARG hits detected in surface water of Haus, Stechlin or Dagow. Considering environmental groups (Fig. 1B-C), the WWTP inflow and outflow accounted for the highest number of classes (i.e., 18 and 16 AMR classes, respectively) followed by urban sediments and urban water (both with 10 classes), rural sediments (6 classes), and farm pond (6 classes each). No AMR classes were detected in the rural surface waters. We could detect up to 511 ARG hits (from a total of 730 hits) in the WWTP inflow and outflow, with the most prevalent drug classes being aminoglycosides (37 hits in the inflow and 31 hits in the outflow), beta-lactams (48 hits inflow and 43 hits in the outflow), biocide resistance (50 hits in the inflow and 31 hits in the outflow) and macrolides-lincosamides-streptogramins (MLS) (42 hits in the inflow and 35 hits in the outflow). A total of 162 ARG hits was predicted in the urban and rural sediments, with the majority of AMR classes associated with aminoglycosides.

**Fig. 1.**
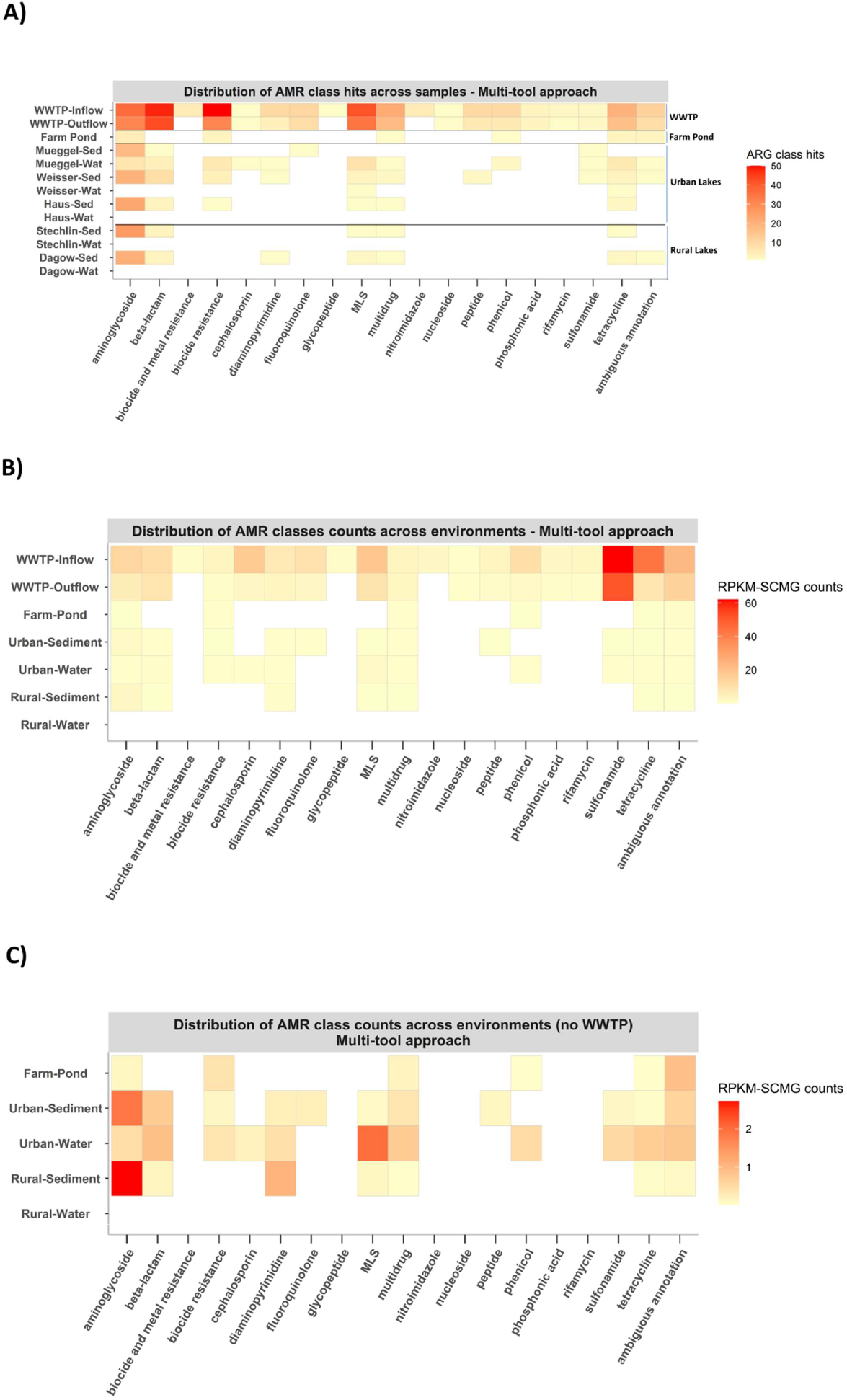
Distribution of antibiotic resistance genes (ARGs) hits and counts predicted in each sample or environmental group and classified by the antibiotic resistance class. Open reading frames (ORFs) were annotated for antibiotic resistance genes (ARGs) and assigned to an AMR class. Each square in the heatmap corresponds to (A) the total number of ARG sequences per sample or (B,C) the mean rank of the reads per kilobase per million mapped reads normalized by single copy marker genes (RPKM-SCMG) linked to an AMR class for each environment, showing (B) or hiding (C) the WWTP. MLS = macrolides-lincosamides-streptogramins.

Both WWTP inflow and outflow had the highest AMR normalized counts (RPKM-SCMG) when compared to all the other environments. The highest abundance was detected in the AMR classes of sulfonamides (in the WWTP inflow and outflow) and tetracyclines (in the WWTP inflow) (Fig. 1B). Other classes that showed high abundance in the WWTP inflow and/or outflow were the aminoglycosides, beta-lactams, cephalosporins, fluoroquinolones, macrolides-lincosamides-streptogramins (MLS) and phenicol. WWTP samples had much higher RPKM-SCMG values than all other samples, therefore we subtracted them to highlight patterns in the other environments (Fig. 1C). Aminoglycosides constitute the AMR class with the highest value of mean RPKM-SCMG counts across all freshwater environments, being more prevalent in the rural sediment than in the urban water. The results show that MLS is more abundant in the urban water compared to the other environments and that urban waters show the highest abundance across all detected AMR classes when compared to other environments. Moreover, diaminopyrimidine is more abundant in rural sediments than in sediments and water from urban environments. We also observed that the farm pond shares most of the predicted AMR classes with urban water, followed by urban sediments.

A total of 6 AMR classes were detected in the sediment samples from rural lakes Dagow and Stechlin, sharing up to six drug classes between the two lakes and differing in two drug classes and diaminopyrimidine (only detected in Dagow) (Fig. 1A). No ARG hits were detected in Haus, Stechlin or Dagow surface water. A higher number of aminoglycoside hits were detected in urban sediments (61 ARG hits) compared to the rural sediments (47 ARG hits). Urban and rural lake sediments exhibited a similar number of aminoglycoside hits, whereas in the water samples, Lake Müggel exhibited the highest number of hits when compared to the other freshwater lakes. The surface water of urban lakes Weißer and Müggel showed differences in the number of AMR classes, with up to 10 AMR classes detected in Müggel and only 2 AMR classes in Weißer (MLS and tetracycline, both shared with Lake Müggel).

### 3.2. Detection profiles of each individual ARG screening tool

Results of the comparison between the detection profile of the multi-tool approach and the ones resulting from using each tool individually are shown in Fig. 2A-F. The maximum number of hits detected per environment and AMR class differed between ARG screening tools, with DeepARG (based on machine learning) revealing the highest number with 972 ARG hits detected, followed by ABRicate with 749 ARG hits, RGI with 578 ARG hits, AMRFinderPlus with 479 ARG hits, and finally Staramr with 264 hits. The multi-tool approach reported a total of 730 ARG hits per AMR class and environment which were often detected by different combinations of tools and databases. There were 11 AMR classes out of 18, i.e. aminoglycosides, beta-lactams, diaminopyrimidine, fluoroquinolones, macrolides-lincosamides-streptogramins (MLS), peptide, phenicol, phosphonic acid, rifamycin, sulfonamide, tetracycline, which were reported by all ARG tools used in this study.

**Fig. 2.**
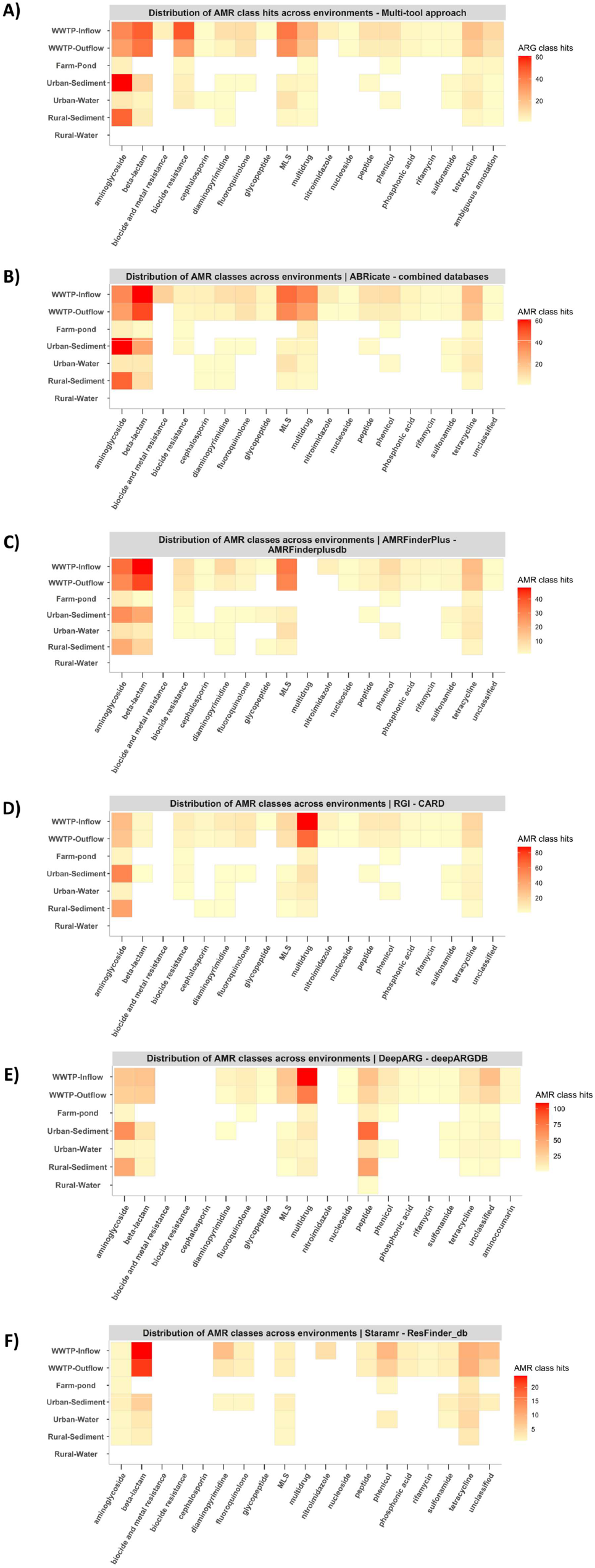
Distribution of ARGs hits predicted in each environment and classified by the antibiotic resistance class for each of the ARG screening tools included in the study. Panels indicate different screening tools (A) multi-tool approach, (B) ABRicate, (C) AMRFinderPlus, (D) RGI, (E) DeepARG, and (F) Staramr. ORFs were annotated for ARGs separately by each ARG screening tool and assigned to a certain AMR class. Each square in the heatmap corresponds to the total number of ARG sequences linked to an AMR class and environment (sample group). The drug class biocide resistance and biocide and metal resistance are part of MEGAres database but is not included in the CARD database.

The tools that performed most similarly to the multi-tool approach (Fig. 2A, Table 1) by detecting a higher number of shared AMR classes were ABRicate (which shared all the 18 AMR classes), followed by RGI (which shared 17 AMR classes), AMRFinderPlus (which shared 16 AMR classes), DeepARG (which shared 15 AMR classes) and Staramr (which shared 12 AMR classes). Both ABRicate and AMRFinderPlus detected a higher prevalence of aminoglycosides, beta-lactams and MLS in the WWTP inflow and outflow and a higher number of aminoglycosides in the urban and rural sediments.

**Table 1.** Detection profile of the 5 ARG screening tools used in the multi-tool approach. The following fields are listed for each tool: Total number of AMR classes detected, Number of detected classes compared to the 18 AMR classes detected by the multi-tool approach, Undetected AMR classes when compared to the multi-tool approach, Total number of ARG hits and Distinct AMR classes detected from the multi-tool approach.

| ARG screening tool | Total drug classes detected | No. of drug classes undetected compared to multi-tool approach | Undetected drug classes compared to multi-tool approach | Total No. of drug resistance hits | Distinct drug classes detected compared to multi-tool approach |
| --- | --- | --- | --- | --- | --- |
| ABRICATE | 18 | 0 | - | 749 | - |
| RGI | 17 | 1 | biocide and metal resistance | 578 | - |
| AMRFinderPlus | 16 | 2 | biocide and metal resistance and multidrug classes | 479 | - |
| DeepARG | 15 | 4 | biocide and metal resistance, biocide resistance, cephalosporin and nitroimidazole | 972 | aminocoumarin |
| Staramr | 12 | 6 | biocide and metal resistance, biocide resistance, cephalosporin, glycopeptide, multidrug and nucleoside | 264 | - |
| ARG screening tool | Total drug classes detected | No. of undetected drug classes compared to individual tools | Undetected drug classes compared to individual tools | Total No. of drug resistance hits | Distinct drug classes detected compared to multi-tool approach |
| Multi-tool approach | 18 | 1 | aminocoumarin | 730 | biocide and metal resistance |

From all 5 tools, only DeepARG detected a different drug resistance class, the aminocoumarin class (in the WWTP inflow, outflow and urban water), which was not reported by the multi-tool approach or any other tools.

### 3.3. Differences in mean ARG counts across different environments

Four AMR classes (aminoglycosides, beta-lactams, macrolides-lincosamides-streptogramins (MLS), tetracycline) and the drug class of biocide resistance showed differences in the mean rank of the RPKM-SCMG counts between different environments, according to the Kruskal-Wallis test, the Dunn’s test for pairwise multiple comparisons and computing Bonferroni adjusted *p-*values (Fig. 3).

**Fig. 3.**
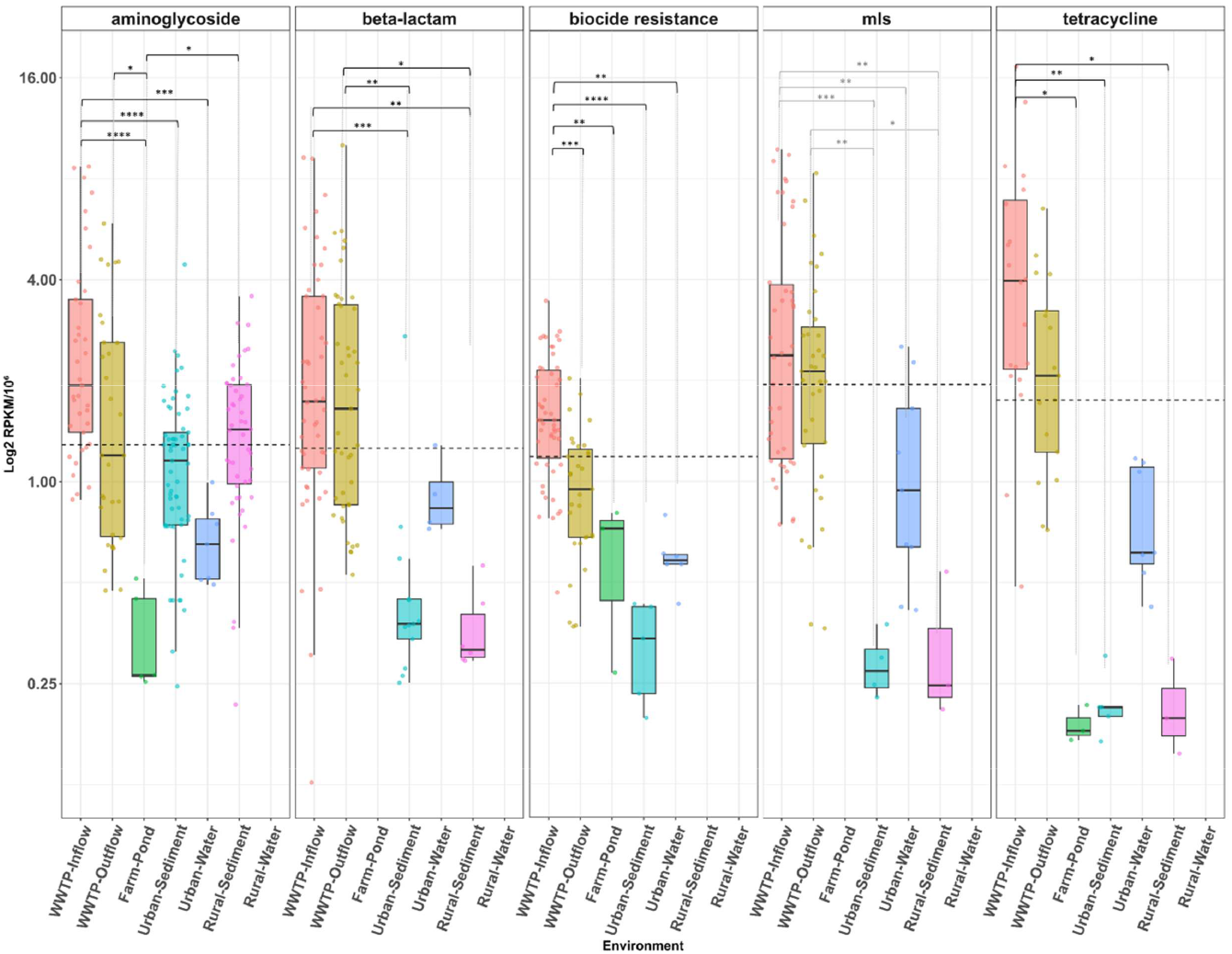
AMR class abundance comparison for those AMR classes that showed significant differences between environments. ORFs were annotated for ARGs separately with the multi-tool approach and assigned to a certain AMR class. The mean rank of the RPKM-SCMG counts was computed by mapping the high-quality reads to the ORFs. A logarithmic scale has been used to represent the RPKM-SCMG counts in the Y axis. The dashed line represents the average of the mean rank of the RPKM-SCMGs counts per each AMR class. P-values significance {^*^ < 0.05, ^**^ < 0.01, ^***^ < 0.001, ^****^ < 0.0001} computed with Dunn test for pairwise multiple-comparison.

For the aminoglycoside AMR class, the mean rank of ARG counts in the WWTP inflow was significantly higher than in all other environments except for WWTP outflow and rural sediments (adjusted *p-*value < 0.001, Table S4, Table S5). The mean rank of aminoglycoside ARG counts was significantly lower in the farm pond when compared to the WWTP inflow (adjusted *p-*value < 0.05, Table S4, Table S5). In the case of beta-lactams, the mean rank of both WWTP inflow and outflow counts were significantly higher than urban (adjusted *p-*value < 0.0001, Table S4, Table S5) and rural sediments (adjusted *p-*value < 0.01, Table S4, Table S5). For MLS, the mean rank of WWTP inflow was significantly higher than urban water (adjusted *p-*value < 0.05, Table S4, Table S5). Another drug class that showed statistical differences amongst environments were genes conferring resistance to biocides, which had a higher mean rank count in WWTP inflow compared to urban water (adjusted *p-*value < 0.001, Table S4, Table S5), WWTP outflow (adjusted *p-*value < 0.01, Table S4, Table S5) and urban sediments (adjusted *p-* value < 0.01, Table S4, Table S5). Finally, for tetracyclines the ARG mean rank count in WWTP inflow was significantly higher than in urban sediments (adjusted *p-*value < 0.01, Table S4, Table S5), farm pond (adjusted *p-*value < 0.05, Table S4, Table S5), and rural sediments (adjusted *p-*value < 0.05, Table S4, Table S5). No ARGs were detected in rural water for any of the AMR classes predicted using the multi-tool approach.

The amount of ARGs shared across urban environments is much higher for WWTP than lakes (Fig. 4 and Fig. S3). In total, 144 ARGs were common between the WWTP inflow and outflow and 70 ARGs were found exclusively in the WWTP inflow, indicating that from a total of 214 ARG hits detected exclusively in the WWTP inflow and outflow, 32.7% of these ARGs were filtered out during the WWTP treatment (Fig. 4 and Table S3). Twenty-three ARG hits were only present in WWTP inflow, outflow, and urban water and 21 ARG hits, mainly associated to aminoglycoside resistance, were found simultaneously between urban and rural sediments. Sixteen ARG hits were shared between urban sediments and WWTP inflow and outflow. The ARGs shared with freshwater environments and WWTP inflow and outflow mostly belonged to the drug classes aminoglycosides, beta-lactams, biocide resistance, MLS, and tetracyclines. Fewer ARG hits that were identified simultaneously in multiple environments were shared between WWTP inflow, outflow and other freshwater environments or the farm pond (Table S6).

**Fig. 4.**
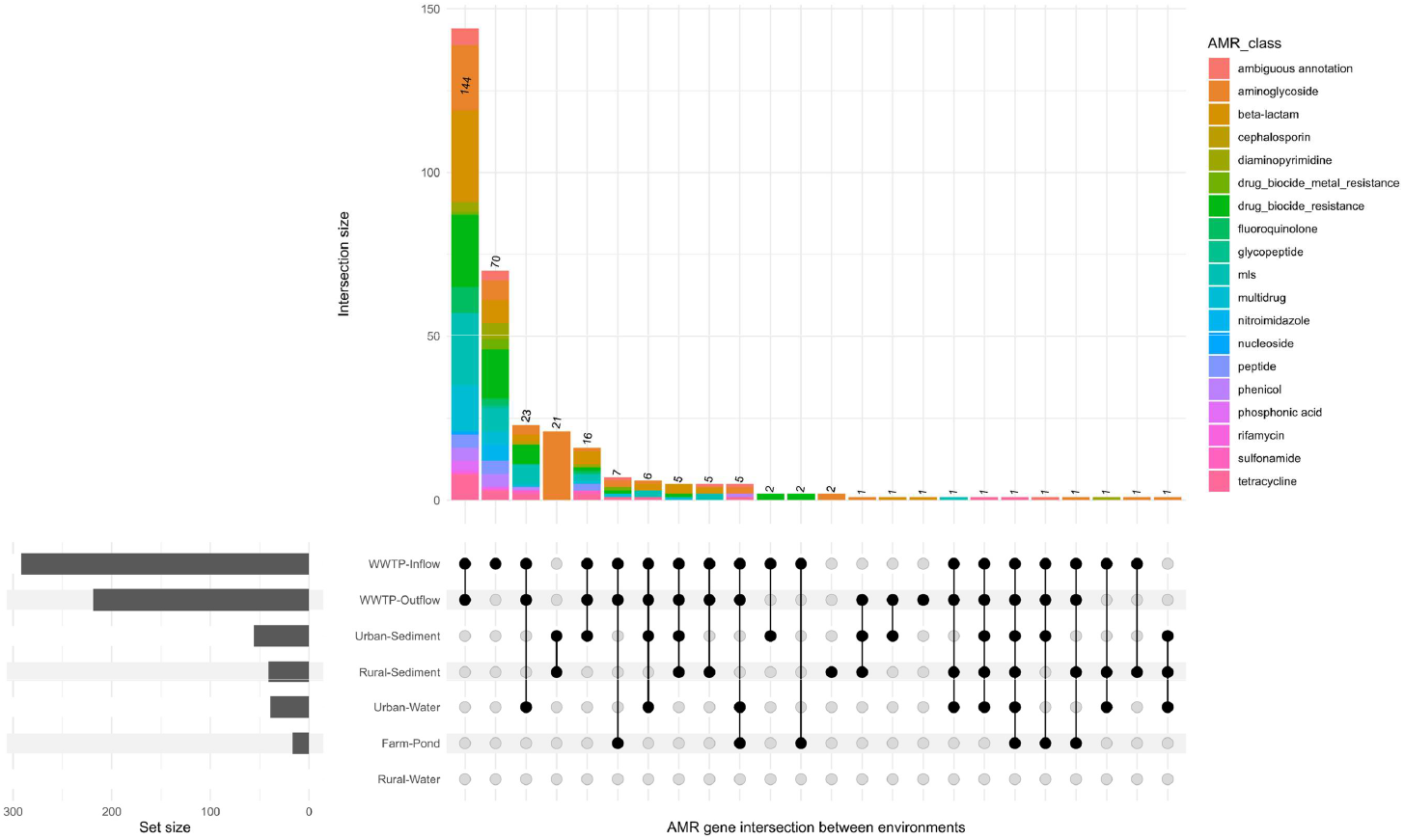
Number of ARGs intersected between the different environments. ORFs which were annotated for ARGs were linked to the different environments by mapping the high-quality reads of each sample back to the ORFs. Each bar in the top panel corresponds to a set of ARGs that are unique to a particular combination of environments (indicated by dot and lines). The number ARGs per each environment is depicted at the bottom left of the plot by horizontal bars.

The *Gammaproteobacteria* was the most predominant taxa detected in all environments, except in rural water where no taxon was annotated (Fig. 5A). WWTP inflow and outflow exhibited higher RPKM-SCMG counts than samples from any of the freshwater environments. Up to 6 different phyla were annotated in WWTP inflow and outflow but there was no annotation for *Cyanobacteria*, which was previously reported in urban and rural sediments. *Gammaproteobacteria* were mainly associated with aminoglycosides, beta-lactams, MLS in both WWTP inflow and outflow and tetracycline in WWTP inflow. Biocide resistance genes were also predicted from WWTP inflow and outflow at levels higher than 4 RPKM-SCMG counts (*1*^*st*^ *Quartile: 0*.*495 RPKM-SCMG, Median: 1*.*67 RPKM-SCMG, 3*^*rd*^ *Quartile: 5 RPKM-SCMG*). Up to 4 different phyla could be detected in the sampled freshwater systems (i.e. *Bacteroidota, Pseudomonadota, Bacillota* and *Cyanobacteriota*). *Cyanobacteriota* could be annotated from rural and urban sediments and urban water showing a similar profile to the farm pond, with *Clostridia* associated with tetracycline. Five taxonomic classes were detected in urban sediments, 4 taxonomic classes in urban water and rural sediments, 3 in the farm pond and no taxonomic classes from any rural surface water samples.

**Fig. 5.**
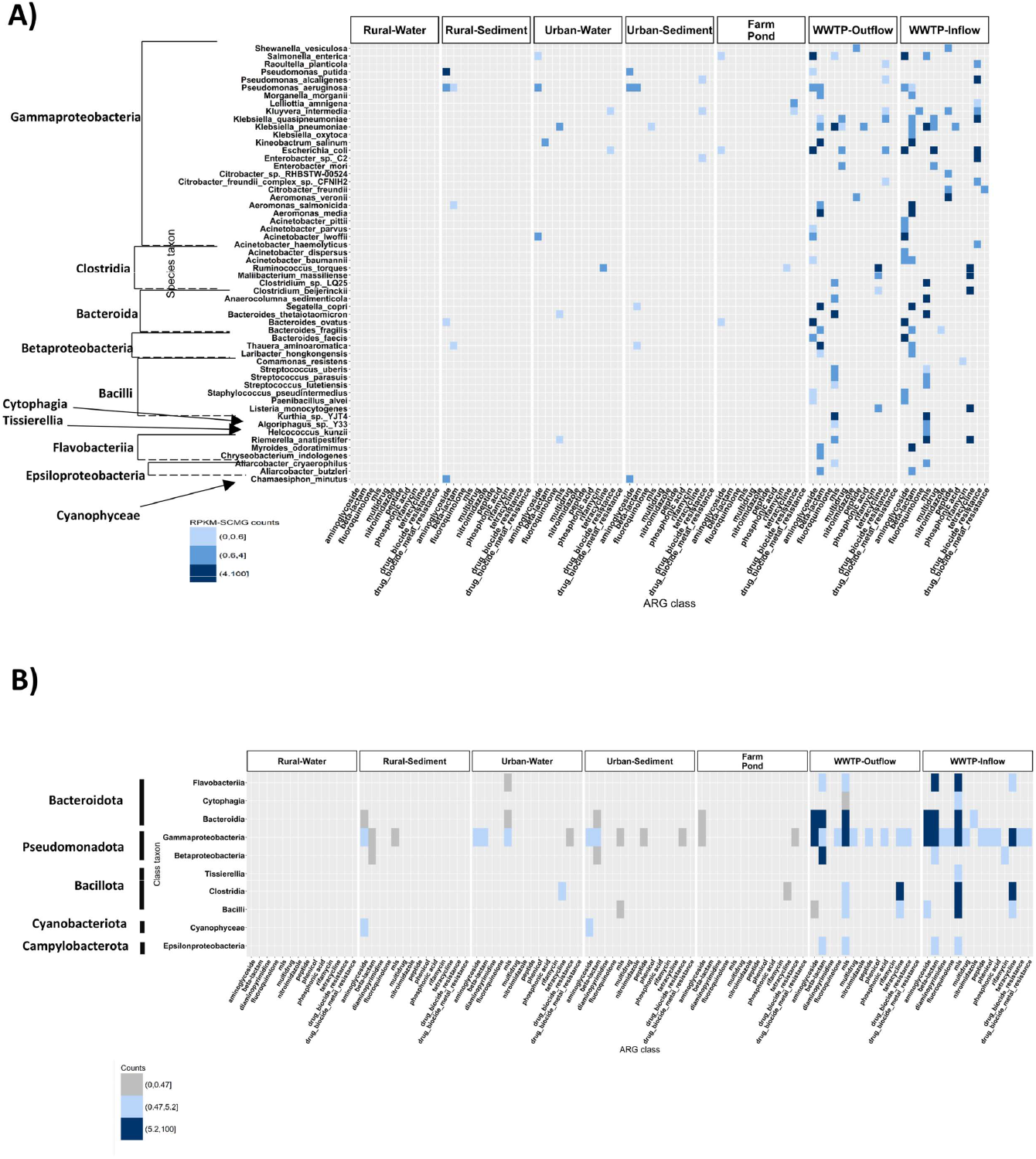
Association between the taxonomic rank and the AMR class annotation in each of the environments. Panels A-B) corresponds to different taxonomic ranks: A) species and B) class level. ORFs were annotated for ARGs and for taxonomic annotation using Kraken2 with the core_nt Database. The mean rank of RPKM-SCMG counts was computed by mapping the high-quality reads to the ORFs. Each square in the heatmap corresponds to the ARG mean rank of the RPKM-SCMG associated to a specific taxon and a specific environment.

At the species level, WWTP inflow and outflow had a higher diversity of species and AMR classes than any other sample type (Fig. 5B). Several species were found to be enriched in both WWTP inflow and outflow, e.g. *Pseudomonas aeruginosa*, which was also reported in rural sediments and urban waters and sediments (associated with aminoglycoside and beta-lactams ARGs), *E. coli* (associated with aminoglycoside ARGs), *Klebsiella pneumoniae* (associated with MLS ARGs), *Ruminococcus torques* (associated with tetracycline ARGs), *Salmonella enterica* (associated with beta-lactam ARGs). For some of the taxa mentioned above, we could detect up to 4 ARGs which were annotated to a particular taxon and environment, potentially indicating the presence of multi-resistant bacteria. In the case of *Pseudomonas aeruginosa*, both aminoglycoside and beta-lactam ARGs were annotated in the rural-sediments, the urban sediments and the WWTP inflow and outflow. Taxonomic annotation indicated that, amongst all the freshwater systems, urban water accounted for the highest diversity of species (10 different taxa) linked to four classes (*Gammaproteobacteria, Clostridia, Bacteroidia* and *Flavobacteria*) and predominantly associated with aminoglycoside and MLS resistance. In the farm pond, rural and urban sediment aminoglycoside was also a prevalent AMR class associated with different classes. For both urban and rural sediments, different species such as *Pseudomonas aeruginosa, Aeromonas salmonicida* or *Thauera aminoaromatica* were associated with the beta-lactam AMR class.

The two most abundant aminoglycoside families in WWTP inflow and outflow were APH(3”) and APH(6), both more abundant in the inflow than in the outflow (Fig. 6A). The farm pond shows a different pattern of aminoglycoside family distribution when compared to all other freshwater environments, sharing the APH(3’) and APH(6) families with WWTP inflow and outflow. The abundance of the aminoglycoside families in the sampled freshwater systems (Fig. 6B) highlights that rural sediments harbour the highest mean RPKM-SCMG counts for AAC(6’), which is the most abundant aminoglycoside family predicted from all sampled freshwater environments. AAC(6’) in urban sediment showed the second highest mean abundance, which was higher when compared to any other mean count from the aminoglycoside gene families predicted from urban water or the farm pond.

**Fig. 6.**
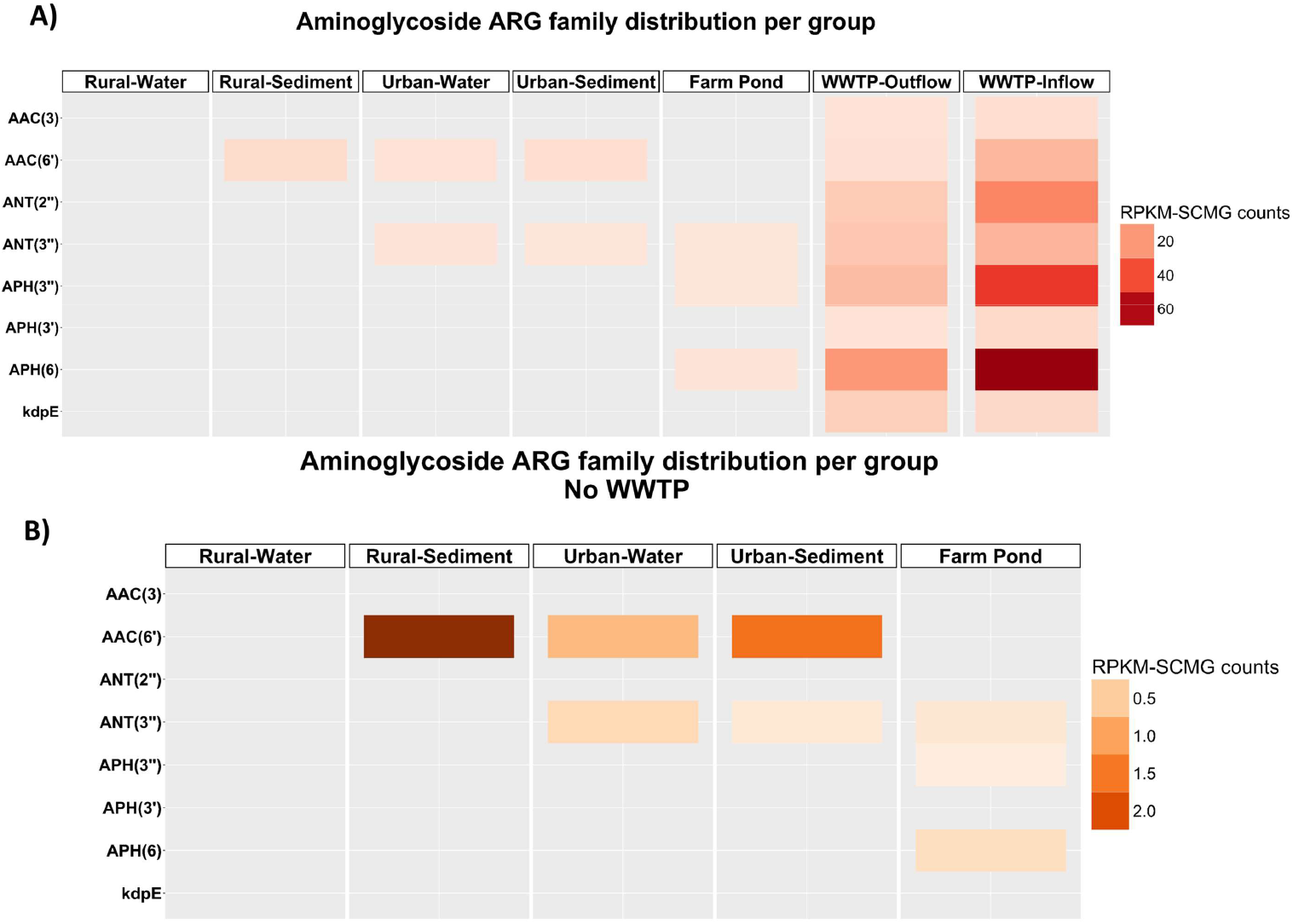
Distribution of aminoglycoside resistance gene families across the different environments ((A) including or (B) excluding the wastewater treatment plant (WWTP). Open reading frames (ORFs) were annotated for antibiotic resistance genes (ARGs), filtered by aminoglycoside class and grouped in aminoglycoside gene families according to CARD database (Alcock et al., 2023). The mean rank of the RPKM-SCMG counts was computed by mapping the high-quality reads to the ORFs. Each square in the heatmap corresponds to mean rank RPKM-SCMG counts associated to an aminoglycoside gene family and environment.

Annotations of ORFs containing plasmid sequences were conducted using geNomad, which can identify and classify plasmids from sequencing data by conducting protein mapping to reference genomes of both isolates and uncultivated species. A total of 3 plasmid sequences could be predicted in 3 ORFs associated with WWTP inflow and outflow or rural sediments (Fig. 7). Out of all the ORFs linked to plasmids, one ORF was annotated as diaminopyrimidine ARG (detected in WWTP inflow and outflow), another ORF as an aminoglycoside ARG (detected in WWTP inflow and outflow) and the last ORF as MLS ARG (detected in WWTP inflow and outflow, and “rural” Lake Stechlin sediments). The aminoglycoside ARG linked to plasmid sequences which was detected in the WWTP inflow and outflow may have been hosted by *Staphylococcus pseudointermedius*, as predicted by geNomad.

**Fig. 7.**
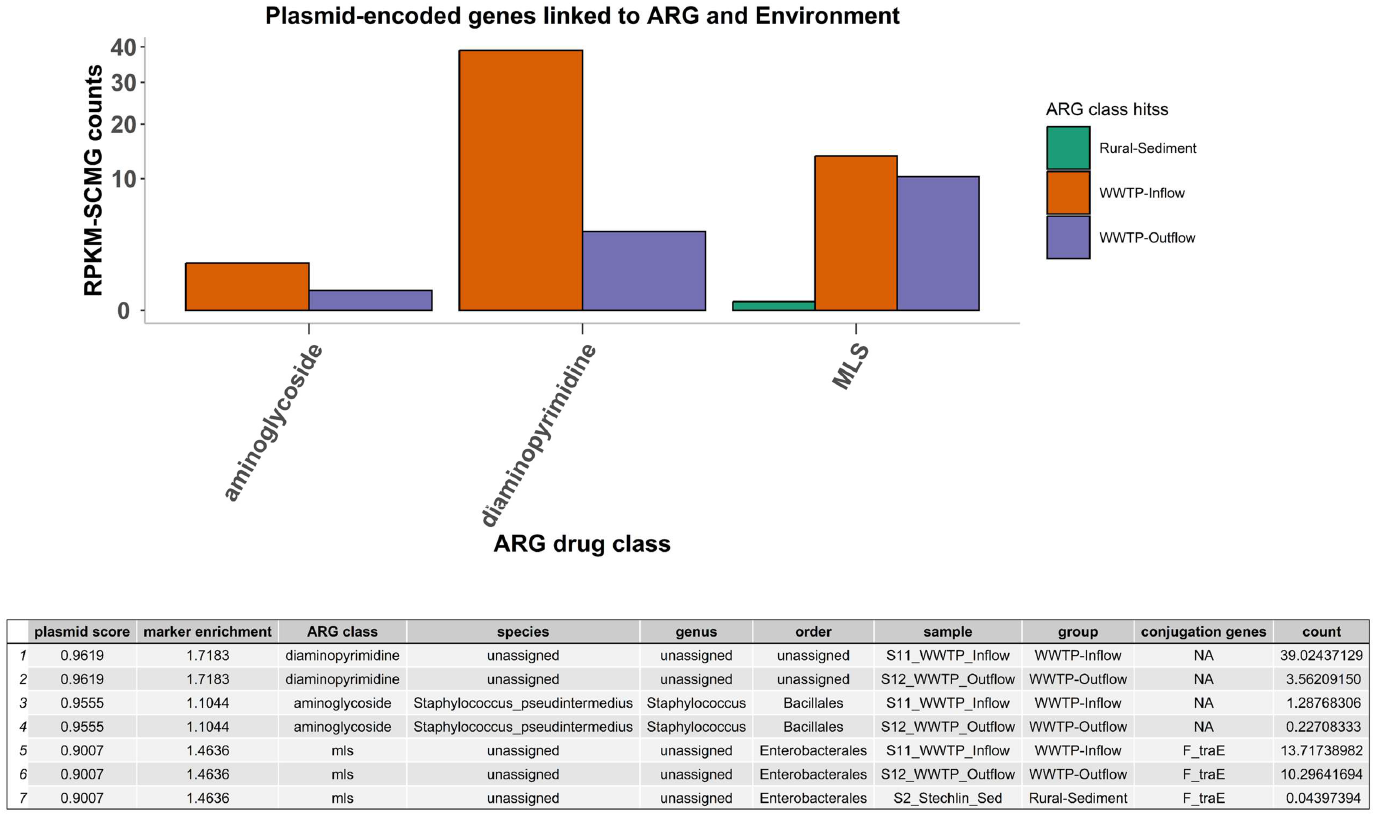
Number and sample group distribution of Open reading frames (ORFs) annotated simultaneously as ARGs and Mobile genetic elements (MGEs). Open reading frames (ORFs) were annotated for antibiotic resistance genes (ARGs) and for mobile genetic elements with geNomad v1.8.1. Only those AMR classes and environments for which at least one hit was present are represented in the figure. The ^*^ indicates that F_traE conjugation genes were detected in the plasmid-encoded genes.

WWTP inflow and outflow showed the highest abundance (RPKM-SCMG) for ARGs linked to plasmid sequences (Fig. 7), like diaminopyrimidine, MLS and aminoglycosides. Furthermore, only the ORF sequence annotated as MLS ARG appears to code for conjugation genes (F_traE). No plasmid sequences were detected in ORFs from urban and rural water or the farm pond. No viral sequences were detected in any ORFs containing ARGs in any of the samples.

## 4. Discussion

### 4.1. The multi-tool approach detects more AMR classes than single-tool approaches

Correct assessment of an ARG tool’s performance relies on determining its prediction accuracy, with low rates of false negatives and false positives. Several studies compared the accuracy of multiple ARG tools based on phenotypic testing or by using ARGs reported in the literature as references (Cooper et al., 2020; Marini et al., 2022; Gomes et al., 2023; Wissel A et al., 2023). Amongst these, some reported minor differences between the tools used, stating that minimal differences were observed when analysing assembled or raw-reads. If the AMR prediction profiles in each study are summarized (Table S2) one can observe that differences in the types of input samples between studies significantly impact the number of AMR classes predicted by the same tool, for example, the number of AMR classes detected with DeepARG in this study was twice the one in (Marini et al., 2022). Other differences can be observed when comparing the performance of one tool among different studies; Gomes et al. (2023) reports that ABRicate predicts the highest AMR class diversity, whereas Wissel et al. (2023b) show that ABRicate prediction capacity is outperformed by many other tools, and Marini et al. (2022), report a higher accuracy of AMR classes by KARGA, a K-mer based tool. However, a reference for ARG detection by independent methods does not exist, the accuracy of each tool against each other has not been assessed comprehensively.

To this end, we implemented a multi-tool approach that addressed annotation discrepancies among tools and databases, and allowed for an increase in the robustness of ARG prediction. Except ABRicate, none of the individual tools included in this analysis could detect as many AMR classes (a total of 18) as the multi-tool approach. Previous studies using other multiple ARG tools and databases have also reported a broader detection of environmental ARGs than a single-tool approach (Goodarzi et al., 2022; Gomes et al., 2023). Single-tool methods are biased toward clinical ARGs (Papp and Solymosi, 2022; Xu et al., 2024) and, while different ARG databases can share multiple ARGs and cover similar AMR classes, others like CARD or ResfinderFG v2.0 databases hold unique sets of ARGs and AMR classes, like quinolones, sulfonamides and trimethoprim. In fact, the broad ARG diversity predicted by ABRicate suggests that, using multiple and diverse databases rather than multiple algorithms, could significantly increase the coverage of AMR diversity detected from the environment.

To ensure a minimum confidence level for ARG prediction, we required that at least two tools share the same AMR class annotation. One potential drawback of the approach is to underestimate the diversity of AMR classes predicted. In our study, one AMR class was discarded from the analysis (aminocoumarin) as the corresponding ARGs were only reported by one tool, DeepARG (see Fig. 2). However, these annotations may not be false positives and could provide relevant information about less well characterized ARGs detectable by only a few specific tools. Such cases should be further investigated in the lab with PCR-based techniques and phenotypic analyses.

It is also important to highlight other limitations which should be considered when comparing ARG profiles. In our analysis, a different sequencing depth achieved between water samples (rural and urban waters, and farm pond) with sediment and wastewater treatment plant (WWTP) samples could be partly responsible for the ARG diversity difference, as shown by the rarefaction analysis (Fig. S1). Nevertheless, a high abundance and diversity of ARGs was observed in urban water, and the comparisons between the WWTP and the sediment samples were statistically significant.

### 4.2. Higher ARG abundance and diversity in urban lake water and sediment

Previous studies have shown higher concentrations of ARGs in surface water compared to lake sediments (Bai et al., 2022; Guo et al., 2022). We could also observe the same pattern for 4 out of 5 drug classes (beta-lactams, macrolides-lincosamides-streptogramins (MLS), tetracyclines, and biocide). In the case of aminoglycoside, the higher abundance in the urban and rural sediments compared to the urban water could be related to the fact that sediments act as an archive and long-term storage for ARGs that were present in the water (Heß et al., 2018). During the 1950s and 1960s, European states approved national regulations for antibiotic use in animals, which may have resulted in a potential environmental contamination of the water with aminoglycoside antibiotics (Castanon, 2007). In addition, soil microbiotas are known for being important producers of aminoglycoside antibiotics, which could result in a higher aminoglycoside resistance accrued in the sediment (Coates et al., 2022). Antibiotic signatures for lake water and sediments have been characterized for multiple lakes in China, showing that in water bodies, sulfonamides and quinolones are the most prevalent AMR classes, whereas in sediments tetracyclines and quinolones are predominant (Liu et al., 2018; Yang et al., 2018). In the current study, a higher abundance of tetracycline resistance was observed in urban surface water when compared to urban sediments and a farm pond, suggesting that antibiotic resistance prevalence may differ according to lake location or their surrounding abiotic and biotic factors, such as the contact with external ARG contamination sources linked to animal husbandry or agricultural practices. However, sulfonamide antibiotics have been widely used for clinical and veterinary uses, and high prevalence of sulfonamide resistance has been associated to wastewater and mobile genetic elements in lakes with high anthropic impact (Stoll et al., 2012; Jiang et al., 2013a; Yang et al., 2017a). In this regard, we detected abundant sulfonamide resistance genes in the WWTP inflow and outflow as well as in water and sediment samples from urban lakes. Instead, no sulfonamide ARGs were detected in samples from rural lakes or the farm pond, supporting the hypothesis of an association of this AMR resistance with the presence and interaction with human populations.

We also observed a higher number of AMR classes in urban (10 classes) compared to rural sediments (6 classes). This difference also held for surface urban water (10 classes) when compared to rural water (i.e. no AMR classes detected). Surface water from urban Müggel accounted for up to 10 different AMR classes, which is much higher than surface water from urban lakes Weißer (2 classes) and Haus (no ARGs detected). Müggel is used for multiple activities that can account for such heterogeneity, namely fishing, shipping, recreation and as a source of drinking water (Behrendt et al., 1990). Although this lake is directly influenced by the Spree River, no other input from wastewater sources has been reported, thus a low AMR class diversity in the surface water was to be expected. In contrast to Müggel and Weißer, sediments from urban lake Haus exhibited a similar AMR class diversity as sediments from rural Dagow and Stechlin. A potential explanation could be the limited urbanization in the proximity of the three lakes. The diversity of ARGs found in the rural sediments (6 classes) could be explained by the fact that the lakes Dagow and Stechlin and their close vicinity were used, during the period 1949 to 1990, for bird/fish farming and mass tourism, respectively (Klapper and Koschel, 1985; Pritchard, 2024). In particular, diaminopyrimidine resistance detected in the sediments from Dagow could be linked to the prior reported use of trimethoprim for duck and carp raising and sewage discard during the period 1949-1990 (Klapper and Koschel, 1985).

Urban sediments and urban waters have higher ARG diversity when compared to rural sediments, and urban water has a higher ARG abundance when compared to urban and rural sediments, except for aminoglycoside. These results are consistent with reports that aquatic urban sediments harbour a higher ARG abundance when compared to rural aquatic environments (Yang et al., 2018; Yang et al., 2020; Na et al., 2021), and that urban water bodies exhibit a higher ARG diversity and abundance when compared to rural waters (Jiang et al., 2013b; Yang et al., 2017b; Huang et al., 2019; Barrantes-Jiménez et al., 2025). However, few studies have simultaneously compared both urban and rural water (Yang et al., 2017b; McInnes et al., 2021; Zhang et al., 2022a; Zhu et al., 2022). Similarly, in the current study, aminoglycosides, beta-lactams, sulfonamides, phenicols, macrolides, lincosamides, peptides were identified in urban lakes, either in water, sediments or both. Aminoglycosides, beta-lactams, MLS and tetracyclines, were also identified in the rural sediments. In contrast, no rifamycin was detected in farm pond, nor urban or rural lakes and in neither water nor sediments.

Aminoglycosides, tetracycline and phenicol ARGs were also detected in the farm pond, consistent with previous studies that reported the same AMR classes in water samples collected from pig farms (Fu et al., 2022). We only detected fluoroquinolone ARGs in WWTP inflow and outflow, and urban sediments. In contrast, tetracycline ARGs were detected in all environments, except in rural surface water. Aminoglycosides, phenicols and tetracyclines, which are all widely used in animal husbandry, were also found in the farm pond sample.

In summary, ARG abundances were significantly different across environments (Fig 1A and Fig 1B), with for example, aminocoumarins in urban sediments having a very low relative abundance compared to most environments (Fig. 1B).

Nearly 69% of all ARGs were unique to WWTP inflow and outflow compared to only 4.1% of the bacterial taxa. Urban water and WWTP shared 16% of the identified ARGs compared to 3.9% of shared bacterial taxa. These two groups alone comprised 85% of the overlapping ARGs but only approximately 8% of the shared bacterial taxa. In contrast to the ARG diversity observed in our study, the majority of the shared bacterial diversity was highest between urban and rural lakes, indicating a decoupling from ARG diversity. One possible explanation could be that most ARG-carrying bacteria are concentrated in water sources derived from hospital effluent and human wastewater, as in most cases bacteria were exposed to antibiotics. The broader diversity of conditions found in urban freshwater lakes (such as diversity of substrates, temperatures, and other biotic and abiotic factors) could lead to a higher diversity of bacterial taxa and functional capabilities. This could explain the slightly greater taxonomic diversity found in bacterial communities from urban lakes when compared to rural ones. Rarefaction curves (Fig. S1), however, appeared to suggest that a higher sequencing depth would be required to fully characterise surface water ARG diversity, as overall DNA retrieval was low compared to sediments.

### 4.3. Abundance and diversity of ARGs do not differ significantly between WWTP inflow and outflow

We observed, as expected, that the WWTP comprises a much larger AMR diversity than other environments, with almost double the number of AMR classes than the environment with the second most diverse AMR classes being the urban water and sediments (10 AMR classes). We detected 18 AMR classes in the WWTP inflow and 16 of them were also found in the WWTP outflow. None of the AMR classes show significant differences in abundance between the WWTP inflow and the WWTP outflow, although more than 50% of ARG hits (76 out of 144) were not detected in the outflow. Nevertheless, these results indicate that the wastewater treatment does not fully remove ARG-containing bacteria despite yielding a large bacterial concentration reduction.

Glycopeptides and nitroimidazole resistance ARGs are two classes that were found in the WWTP inflow but not in the outflow (as shown in Fig. 1B and Table S7). A possible explanation is that bacteria carrying these ARGs in the WWTP inflow were removed or diluted after the WWTP treatment. Although some specific AMR classes are reported to be largely removed during the wastewater treatment (Tran et al., 2016), WWTPs cannot differentiate or selectively filter out specific ARGs, and detection may be related to the initial abundance of a specific AMR class carried by specific bacteria. Glycopeptide and nitroimidazole consumption in German medical facilities has been reported to be lower than other antibiotics (Schweickert et al., 2018). This could be linked to a decrease of C. difficile infections in Germany (German Federal Institute for Risk Assessment, 2023) together with stricter regulations posed by the European Commission (Polzer et al., 2000). Low prevalence of these antibiotics in the environment could have resulted in low antibiotic resistance detection in the WWTP inflow, which at the same time might have led to no detection in the WWTP outflow.

Our results show that WWTP inflow and outflow have the highest ARG overlap (Fig. 4) with 144 genes shared between inflow and outflow. However, WWTP inflow contained 70 unique ARGs, whereas outflow only contained 1 unique ARG. The fact that the WWTP inflow shows a higher number of unique ARGs may be explained by the fact that wastewater treatment affects the bacterial community in the outflow (Numberger et al., 2022). The removal of bacterial cells, taxa and associated ARGs during the treatment process may lead to ARG levels below the detection limit, even when using deep sequencing techniques. The reduction in ARG abundance in the WWTP outflow compared to the inflow is consistent with previous studies (Yang et al., 2014; Ng et al., 2019).

### 4.4. Microbial community signatures differ between urban and rural environments

The overall distribution of bacterial taxa in each of the different samples is consistent with our previous study using non-pooled samples (Numberger et al., 2022). In the present study we pooled three samples per site and performed metagenomics shotgun sequencing using Illumina short reads. This pooling of samples may explain some of the discrepancies between the previous and the current analysis, particularly for bacterial taxa at low abundance which is easier to detect by PCR-based approaches.

Multiple associations between AMR classes and bacterial taxa could be established at the ORF level. Among all drug classes, aminoglycosides, macrolides-lincosamides-streptogramins (MLS), beta-lactams or tetracyclines predominated and were associated with different bacterial taxa. Most AMR classes were enriched in WWTP inflow and outflow but were also found in urban and rural sediments and the farm pond. We could also observe that these drug classes are linked to specific pathogens as reported by other studies (Garba et al., 2023; Abdulkadir et al., 2024). For example, *Klebsiella pneumoniae* (beta-lactams, MLS, multidrug and phosphonic acid ARGs) appeared exclusively in WWTP inflow and outflow. *Pseudomonas aeruginosa* in the rural sediments was associated with aminoglycoside and beta-lactam.

Three different ORFs potentially linked to plasmid sequences could also be associated with three ARGs, i.e. aminoglycosides, MLS and diaminopyrimidine in WWTP inflow and outflow, with the MLS plasmid also found in rural sediment was the only case where the conjugation genes F_traE were observed. Both aminoglycoside and MLS ARGs have been previously associated with plasmids sequenced from heavily contaminated riverine waters and sediments in Costa Rica (Barrantes-Jiménez et al., 2025). F_traE genes are thought to be involved in the resolution of plasmid DNA replication intermediates (Li et al., 1997; European Medicines Agency, 2018).

### 4.5. Distribution of aminoglycoside in urban and rural environments

Two aminoglycosides ARG gene families could be detected in both urban water and sediments, namely *ANT(3”)* and *AAC(6’)*, the latter also found in rural sediments. *ANT(3”)* is the most common class of *ANTs* enzymes which are ubiquitous across different biomes, including WWTP, freshwater aquatic systems, pig farms as well as human samples (Yang et al., 2009; Kaushik et al., 2019; Shin et al., 2023; Zhou et al., 2023; Makowska-Zawierucha et al., 2025). Two aminoglycoside resistance genes, i.e. *AAC(6’)-IIa* and *AAC(6’)-32*, were also found in rural and urban lake sediments, and *AAC(6’)-II* was found in rural and urban sediments as well as in urban water (Fig. S4). Other aminoglycoside ARGs genes like *AAC(6’)-I, AAC(6’)-Ib* and *AAC(6’)-IE* could not be detected in the rural sediments.

The widespread use of aminoglycoside antibiotics for clinical treatment of human infections is associated with an increase in the abundance of these ARGs in municipal wastewater. Aminoglycoside antibiotics are commonly used to prevent bacterial diseases in animal husbandry, apiculture and aquaculture (European Medicines Agency, 2017; Nowacka-Kozak et al., 2023). The fact that most aminoglycoside ARGs in urban and rural lakes are also reported in wastewater, indicates that aminoglycoside antibiotics application in clinical settings and/or food production tend to enrich a similar pattern of aminoglycoside ARGs in the aquatic environment (Van Duijkeren et al., 2019a).

WWTPs collect and concentrate ARGs from multiple and different urban water sources. In contrast, the farm pond was not connected to any WWTP. However, three aminoglycoside gene families were identified, more than found in other freshwater environments but less than in WWTP inflow and outflow. Aminoglycoside antibiotics, such as gentamicin and tobramycin, are commonly used in German neonatal intensive care units (NICUs) to treat Gram-negative infections. At the same time, in 2015, the sales of aminoglycosides when aggregating 30 European countries made up for 3.5% of the total sales of antimicrobials for food production, and the sixth most used antimicrobial class for veterinary medicine (European Medicines Agency, 2015; Van Duijkeren et al., 2019b; Marissen et al., 2021). This may explain the relatively high overlap of aminoglycoside ARGs between the WWTPs and farm pond (3 gene families shared) when compared to WWTP vs. lake water and sediments (2 gene families shared).

## 5. Conclusions

The current study highlights the importance of using a multiple ARG screening approach to detect ARGs in aquatic environments. WWTPs clearly have the highest level of AMR classes, in comparison to urban and rural environments, and could act at times as a potential source of AMR inflow. The results of the current study support the central role of sediments, particularly those in the more contaminated settings (here WWTPs), as an important reservoir for ARGs and showcased the high number of AMR classes detected in both urban and rural lake sediments. The current study is to the best of our knowledge the first to report an ORF sequence linked to F_traE conjugation genes associated with MLS in a natural aquatic habitat. That conjugation genes could be detected in this ORF and not in others suggests a larger distribution of this plasmid (not only in WWTP inflow and outflow water but also in rural sediments) when compared to the occurrence of the other two detected ORFs linked to plasmid sequences (exclusively associated to WWTP inflow and outflow).

Although validation using methods such as PCR-based assays remain essential, future enhancements of databases that better capture environmental AMR diversity will improve precision and accuracy of deep sequencing-based approaches, ultimately strengthening environmental AMR surveillance.

## Data Availability

All data produced in the present study are available upon reasonable request to the authors

## Resource availability

### Lead contact

Further information and requests should be directed to and will be fulfilled by the corresponding authors in this journal, Prof. Alex Greenwood and Prof. Hans-Peter Grossart.

## Data and code availability

- This study did not generate any new unique code.
- Raw WGS sequencing data was deposited into the European Nucleotide Archive (ENA), under the accession number PRJEB98077.
- Any additional information regarding the data reported in this publication will be made available by the corresponding author upon request.

## Acknowledgments

We thank Danny Ionescu, Jason Woodhouse, Guilherme Neumann, Dorina Meneghini and Rafael Cuadrat for their computational assistance in this study. We thank Karin Hönig and Solvig Pinnow for technical support. We thank Jaffer Dar, Sabreen Samuel Ibrahim Dawoud and Richard Mugani for their support in the sampling and other technical help. PDY, ADG and HPG were supported by a project grant (IRG 3 - Water) from the Leibniz Research Alliance “INFECTIONS’21 - INFECTIONS in an Urbanizing World - Humans, Animals, Environments” funded by the Leibniz Association, Germany (SAS-2015-FZB LFV).

## Data statement

All data generated in this study will be available upon request.

