## Supplemental methods, figures and tables for "Diversity of antibiotic resistance genes increases in urbanized lakes: a multi-tool screening"

***Correspondence:**

#### 1.1. Sample collection

Lakes Müggel and Weißer in Berlin (the capital of Germany with an area of 891.1 km2 and 3.7 Mio inhabitants) and Lake Haus (located in the small city of Feldberg in Mecklenburg-Vorpommern, in the state of Mecklenburg-Vorpommern) were exposed to pronounced anthropogenic impacts due to previous wastewater input (Krienitz et al., 1996), making them comparable to lakes in bigger cities and are thus defined as urban lakes. None of the urban lakes received direct wastewater input during the sampling period. The two rural lakes, Lake Dagow and Lake Stechlin are located in a forested nature reserve in Northern Brandenburg and have little anthropogenic impact, surrounded by only 383 inhabitants in the villages of Dagow and Neuglobsow. All lakes originate from the last ice age but greatly vary in their present environmental status.

Untreated raw inflow water and treated outflow were sampled from a municipal wastewater treatment plant (WWTP) processing the waste of *ca.* 3.5 million citizens of Berlin (Germany) in 2016 (Amt für Statistik Berlin-Brandenburg, 2017). This WWTP processes a negligible amount of industrial wastewater. Samples were collected at the inflow and the outflow of the WWTP. A water sample from a farm pond was collected in the municipality of GroßKreutz, which is located in the district Potsdam-Mittelmark (Brandenburg, Germany) and has a population of 8,948 inhabitants. The farm pond is located 6 km away from a wastewater treatment plant and more than 1 km distance from Grosskreutz itself. The characteristics of all five lakes, the wastewater treatment plant, and the farm pond are shown in Table S1.

Wastewater inflow and outflow, lake surface water, and sediment samples were collected every three months in 2016 from two locations in Lake Weißer and three different locations in lakes Müggel, Haus, Dagow and Stechlin. Another set of water samples was collected from a farm pond (GrosßKreutz). Further details on sample collection are described in Numberger et al. (2022).

Samples were grouped as follows:

1. Rural-Water and Rural-Sediments (water or sediments from lakes Dagow and Stechlin), 2. Urban-Water and Urban-Sediments (water or sediments from lakes Haus, Müggel and Lake Weißer), 3. Water from a farm pond in GroßKreutz and 4. WWTP (inflow or outflow).

#### 1.2. Illumina short-read sequencing

DNA from the water was extracted from Sterivex® filters (EMD Millipore, Darmstadt, Germany) using the QIAamp DNA mini kit (Qiagen, Hilden, Germany) following the protocol for tissues with some modifications. WWTP and sediment samples showed both a similar semi-solid nature, hence, the DNA from both environments was extracted using the NucleoSpin® Soil kit REF 740472.50 (Macherey 180 Nagel, Düren, Germany), which is suitable for sediment, solid and sludge samples. More details on sample collection are given in Numberger et al. (2022). Replicates from each site (considering WWTP inflow and outflow as separate sites) were pooled to reduce the impact of spatial and temporal heterogeneity. Subsequently, the DNA was sequenced using Illumina PE150 HiSeq X, which resulted in sequencing depths per sample ranging from 5 to 390 million reads. The assessment of the impact of the differences (Fig. S1) showed a high ARG coverage for sediment and WWTP samples but a potential relative underestimation of ARGs from the water samples.

##### 1.3. AMR tools database references

Abricate v 1.0.1

NCBI AMRFinderPlus - doi: 10.1128/AAC.00483-19 (2631 sequences - 2021-Mar-27),

CARD - doi:10.1093/nar/gkw1004 (2631 sequences - 2021-Mar-27),

Resfinder - doi:10.1093/jac/dks261 (3077 sequences - 2021-Mar-27),

ARG-ANNOT - doi:10.1128/AAC.01310-13 (2223 sequences - 2021-Mar-27),

PlasmidFinder - doi:10.1128/AAC.02412-14 (460 sequences - 2021-Mar-27),

MEGARES 2.00 - doi:10.1093/nar/gkz1010 (6635 sequences - 2021-Mar-27)

Amrfinder v 3.11.4

Database version 2023-02-23.1

Deeparg

Deeparg_db version 1.0

Staramr 0.9.1

https://bitbucket.org/genomicepidemiology/resfinder_db/src/master/

Resistance Gene Identifier

rgi database version 3.2.9 (9610 sequences)

KARGA

megares_database_v3.00.fasta and kargva_db_v5.fasta

KARGVA

kargva_db_v5.fasta

**1.4. Steps followed to homogenize the ARG nomenclature**

We renamed those ARG class entries that corresponded to the same drug class but were written differently (e.g., “Aminoglycosides” and “aminoglycoside”, “Multi-drug_resistance” and “multidrug” or “mls” and “lincosamide/macrolide/streptogramin”

To assign the annotation, we considered all the databases used by the tools in this study (CARD, KARGVAdefault, AMRFinderplusdb, MEGARes, ARG-ANNOT, NCBI, ResFinder and DeepARGDB).

The predicted ARGs were annotated based on the following criteria:

- If one annotation was dominant compared to the others we used the most prevalent AMR class to classify the gene (eg. Tetracyclines has an absolute frequency of four and Aminoglycosides of two, we assign the AMR hit as tetracycline).
- In the case, two different annotations were both the two most prevalent ones:
  - If the most prevalent labels were {*two different arg_classes* different from *multidrug or unclassified*} then we added the label "*ambiguous annotation*"
  - If the most prevalent labels were {*multidrug* and another *arg_class* (which is not *multidrug*)} then we added the label of the *arg_class*
  - If the most prevalent labels were {*multidrug* and two other *arg_classes*} then we added the label "*multidrug*"
  - If the most prevalent labels were {*multidrug* and *unclassified*} then we added the label as "*unclassified*"
  - If the most prevalent labels were {*unclassified* and another *arg_class* (which is not *multidrug*)} then we added the label of the *arg_class*

In addition, the following replacements in annotations were conducted according to the CARD database (Alcock, B. P., et al., 2023. CARD 2023: expanded curation, support for machine learning, and resistome prediction at the Comprehensive Antibiotic Resistance Database. Nucleic acids research, 51(D1), D690-D699.):

| **Old label** | **New label** |
|---|---|
| Aminoglycosides | aminoglycoside |
| Tetracyclines | tetracycline |
| aminoglycoside:aminocoumarin | aminocoumarin |
| Aminocoumarins | aminocoumarin |
| Lipopeptides | lipopeptide |
| Multi-drug_resistance | multidrug |
| Sulfonamides | sulfonamide |
| Tetracyclines | tetracycline |
| betalactams | beta-lactam |
| macrolide | MLS |
| LINCOSAMIDE/MACROLIDE/STREPTOGRAMIN | MLS |
| LINCOSAMIDE;MACROLIDE;STREPTOGRAMIN | MLS |
| cationic_antimicrobial_peptides | peptide |
| metronidazole | nitroimidazole |
| chloramphenicol | phenicol |
| rifampin | rifamycin |
| fosfomycin | phosphonic acid |
| streptomycin | aminoglycoside |
| bacitracin | peptide |
| lincosamide | MLS |
| streptothricin | nucleoside |

| | |
|---|---|
| colistin | peptide |
| thiopeptides | peptide |
| elfamycins | elfamycin |
| fluoroquinolones | fluoroquinolone |
| fosmidomycin | phosphonic acid |
| fusidic_acid | fusidane |
| glycopeptides | glycopeptide |
| lipopetide | peptide |
| metronidazole | nitroimidazole |
| nucleosides | nucleoside |
| rifampin | rifamycin |
| lincosamide/streptogramin | MLS |
| phenicol/quinolone | unclassified |
| quinolone | fluoroquinolone |
| lincosamide | MLS |
| fosfomycin | phosphonic acid |
| colistin | peptide |
| fluoroquinolone | quinolone |
| glycopeptide | glycopeptides |
| macrolide | MLS |
| drug_biocide_resistance | biocide resistance |
| drug_biocide_metal_resistance | biocide and metal resistance |
| mls | MLS |
| trimethoprim | diaminopyrimidine |
| ammonium | biocide resistance |
| cephamycin | beta lactam |

### Supplementary figures

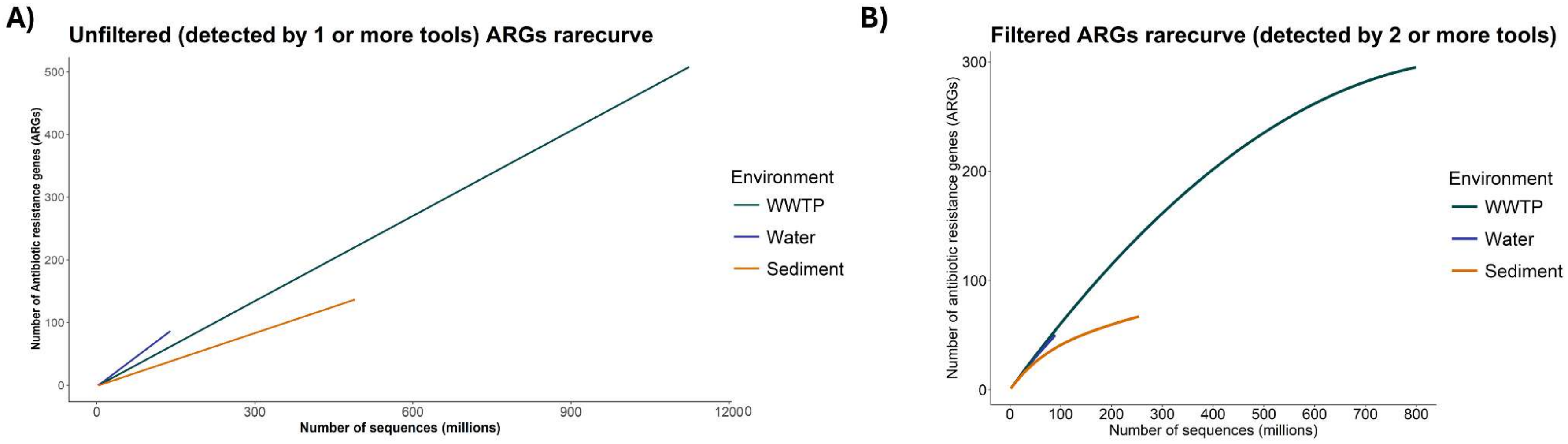


**Fig. S1.- Rarefaction curves for the coverage ARGs in relationship to the sequencing depth.** After annotating the ORFs for ARGs, each ARG was assigned to one or more environments. Rarefaction curves were computed by grouping the ARGs into three sample types: water, sediments and WWTP. Panel A) uses as input the unfiltered ARG dataset (all the ARGs predicted by at least 1 screening tools) and B) uses as input the filtered dataset (only those ARGs predicted by at least 2 screening tools). The rarefaction was computed using the function rarecurve from the package v*egan* version 2.6-6.1

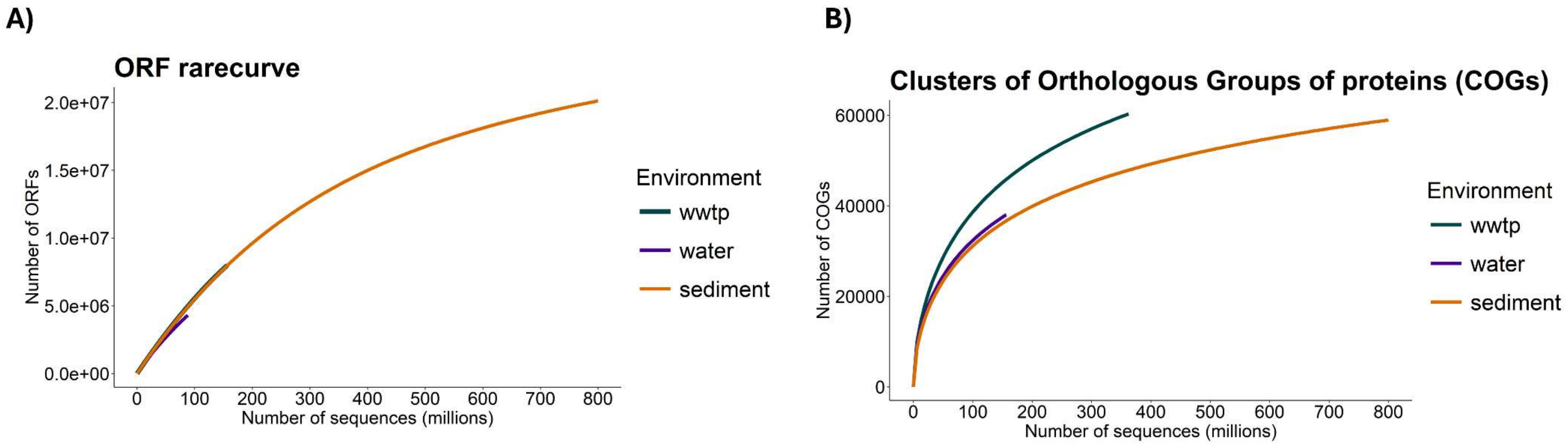


**Fig. S2.- Rarefaction curves for the coverage of (A) ORFs and (B) Clusters of Orthologous Groups of proteins (COGs) in relationship to the sequencing depth.** ORFs were annotated for COGs using eggnog mapper v2.1.5. Rarefaction curves were computed by grouping the ORFs and COGs into three sample types: water, sediments and WWTP. The rarefaction was computed using the function rarecurve from the package v*egan* version 2.6-6.1

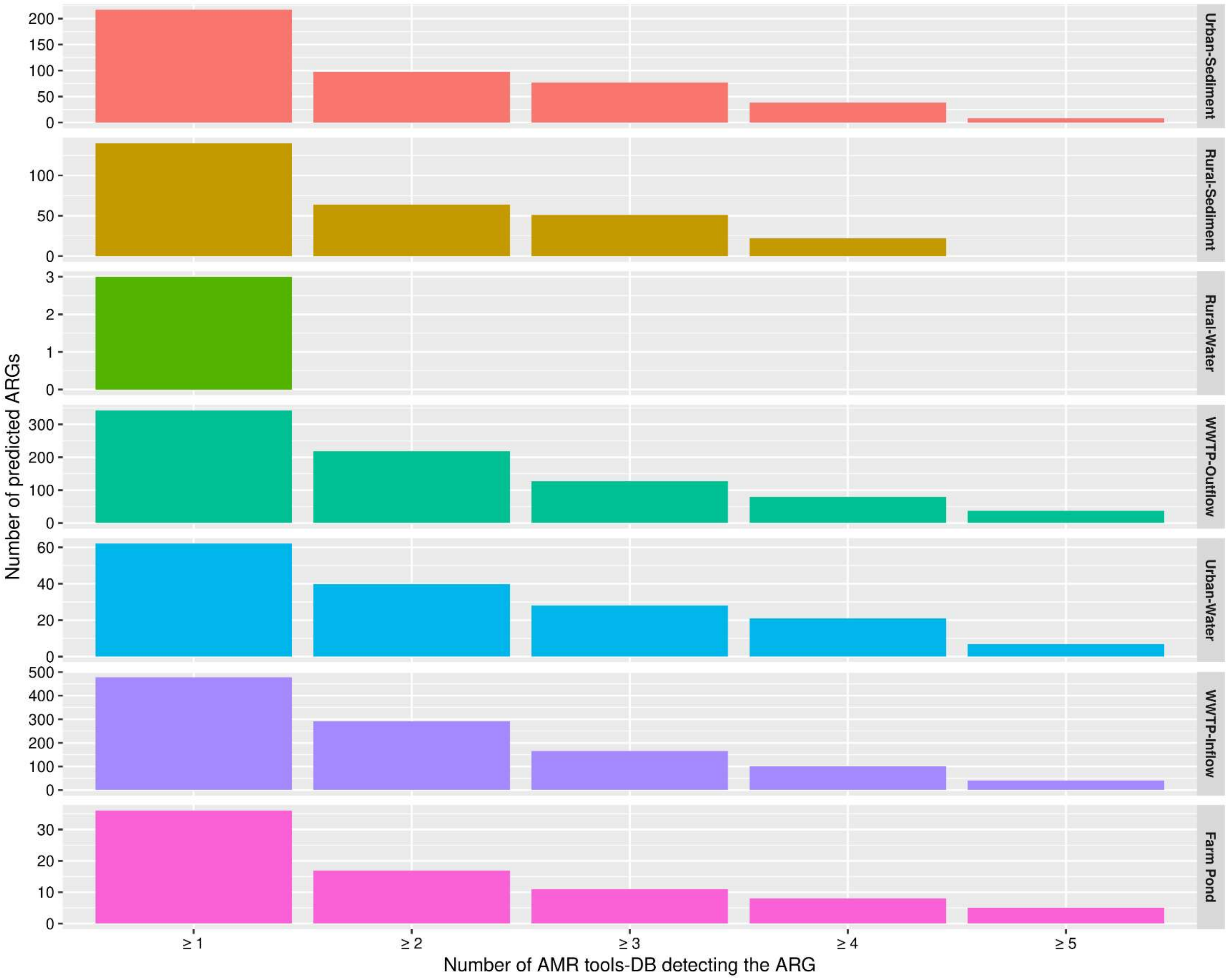

Number of predicted ARGs
Number of AMR tools-DB detecting the ARG
≥ 1
≥ 2
≥ 3
≥ 4
≥ 5
Urban-Sediment
Rural-Sediment
Rural-Water
WWTP-Outflow
Urban-Water
WWTP-Inflow
Farm Pond

**Fig. S3. - Number of ARGs hits per environment that were predicted by the multi-tool approach based on the number of ARGs screening tools that detected each ARG.** ORFs were annotated ARGs using 5 ARG tools and then merged together. Then each ARG was grouped depending on how ARG tools annotated the ARG. The first column corresponds to those ARGs detected by at least one tool whereas the last column contains those ARGs detected by all 5 tools.

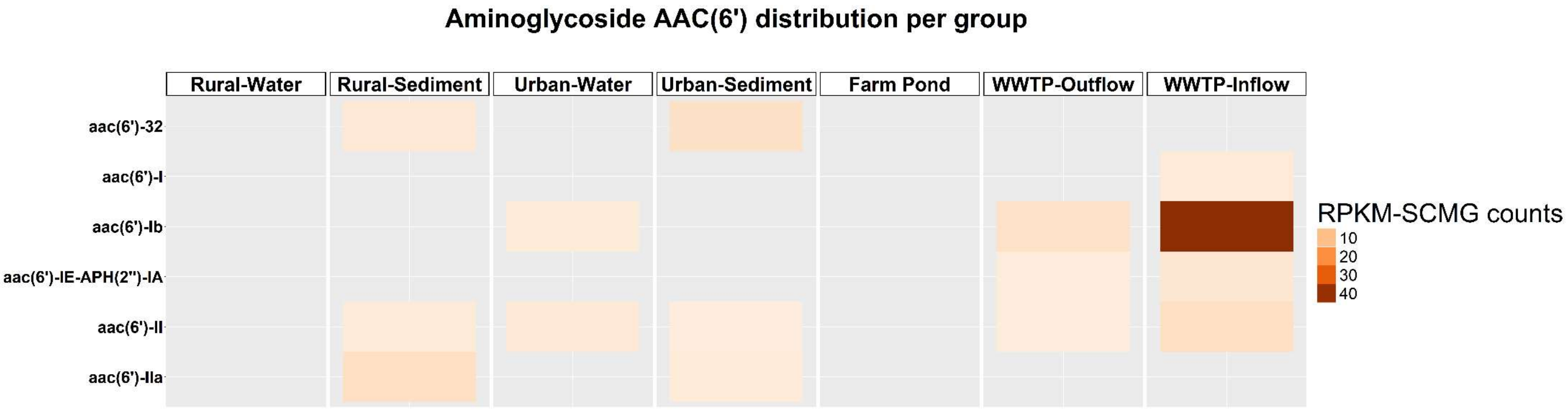


**Fig. S4.- Distribution of aminoglycoside gene family AAC(6') across the different environments.** Open reading frames (ORFs) were annotated for antibiotic resistance genes (ARGs), filtered by aminoglycoside class and grouped in aminoglycoside gene families according to CARD database (Alcock et al., 2023). Only those genes belonging to the aminoglycoside family AAC(6') were selected. The mean rank of the reads per kilobase per million mapped reads normalized by single copy marker genes (RPKM-SCMG) counts was computed by mapping the high-quality reads to the

ORFs. Each square in the heatmap corresponds to mean rank RPKM-SCMG counts associated to an aminoglycoside AAC(6') gene family and a particular environment.

#### Supplementary table

**Table S1. Sampling sites and their characteristics.** Samples are grouped in four groups: Wastewater treatment plant (WWTP) inflow and outflow; Urban waters which include the lakes, 'Lake Feldberger Haussee', 'Lake Müggelsee' and 'Lake Weißer See'; Rural lakes which include 'Lake Stechlin' and 'Lake Dagow'; and farm pond with a water sample from a farm in Groß Kreutz (Brandenburg). The following fields are listed for each sample: Geographical position, Catchment Area, Inhabitants in the catchment area, Maximum depth (for WWTP is the treatment of wastewater per year), Trophy status and cited References. The exact location of the sampled WWTP cannot be disclosed due to a confidentiality agreement with the WWTP operators.

| Water | Geographical position | Catchment Area | Inhabitants in catchment area | Max. Depth [m] | Surface area [$km^2$] | Trophy | Ref |
|---|---|---|---|---|---|---|---|
| Wastewater Treatment Plant | Confidential, in agreement with WWTP | 1.6 million inhabitants (of Berlin) | | Treatment of 247,500 $m^3$ raw wastewater per day | | Hyper-eutrophic | 1 |

| | operators | | | | | | |
|---|---|---|---|---|---|---|---|
| Müggelsee | 52°26'N, 13°39'O | Berlin Treptow-Köpenick | 271,153 (1,610/km²) | 8 | 7.3 | eutrophic | 2, 3 |
| Weißer See | 52°33'N, 13°27'O | Berlin Weißensee | 53,737 (6,776/km²) | 10.6 | 0.08 | eutrophic | 4 |
| Feldberger Haussee | 53°20'N, 13°26'O | Feldberger Seenlandschaft | 4,433 (22/km²) | 12 | 1.3 | eutrophic | 5, 6 |
| Dagowsee | 53°09'N, 13°03'E | Dagow and Neuglobsow | 383 | 9.5 | 0.3 | eutrophic | 7–9 |
| Stechlinsee | 53°10'N, 13°02'E | | | 69.5 | 4.3 | oligo-meso-trophic | 10, 11 |
| Großkreutz | 52°23'47.3"N, 12°45'57.9"E | Potsdam-Mittelmark | 222,570 | - | - | - | - |

**Table S2. Comparison of the AMR class prediction (type and number of AMR classes) reported by this and previous studies when using multiple AMR screening tools and databases.** Each row corresponds to a single combination of AMR screening tool and database. For each combination the following fields regarding the AMR prediction capacity are listed: AMR tool, Database, Drug classes detected, Number of drug classes detected, Sample type and Reference. The "*" indicates that those predictions showed a very low Balanced Accuracy (BalAcc), near 0.50, resulting in an AMR class detection which is likely to be inaccurate (Marini et al., 2022).

| | AMR Tool | Database used | Drug classes detected | Number of drug classes detected | Input sample | Reference |
|---|---|---|---|---|---|---|
| 1 | **Multi-tool approach** (Same tools from rows 2-6, filtering hits detected by ≥ 2 tools) | Same databases from rows 2-6 | Aminoglycosides, Beta-lactams, Cephalosporins, Diaminopyrimidine, Fluoroquinolone, Glycopeptide, MLS, Multidrug, Nitromidazole, Nucleoside, Peptide, Phenicol, Phosphonic acid, Rifamycin, Sulfonamide and Tetracycline. Additionally drug resistance against Biocides and Metals. | 18 | Detection in at least one environment amongst WWTP inflow, WWTP outflow, farm pond or freshwater lakes (water or sediments) | This study |

| | | | | | | |
|---|---|---|---|---|---|---|
| 1 | **ABRICATE** | ResFinder DB, NCBI, ARG-ANNOT, CARD and MEGARes | Aminoglycosides, Beta-lactams, Cephalosporins, Diaminopyrimidine, Fluoroquinolone, Glycopeptide, MLS, Multidrug, Nitromidazole, Nucleoside, Peptide, Phenicol, Phosphonic acid, Rifamycin, Sulfonamide and Tetracycline. Additionally drug resistance against Biocides. | 18 | Detection in at least one environment amongst WWTP inflow, WWTP outflow, farm pond or freshwater lakes (water or sediments) | This study |
| 2 | **RGI** | CARD | Aminoglycosides, Beta-lactams, Cephalosporins, Diaminopyrimidine, Fluoroquinolone, Glycopeptide, MLS, Multidrug, Nitromidazole, Nucleoside, Peptide, Phenicol, Phosphonic acid, Rifamycin, Sulfonamide and Tetracycline. Additionally drug resistance against Biocides. | 17 | Detection in at least one environment amongst WWTP inflow, WWTP outflow, farm pond or freshwater lakes (water or sediments) | This study |
| 3 | **AMRfinderplus** | AMRfinderplus DB | Aminoglycosides, Beta-lactams, Cephalosporins, Diaminopyrimidine, Fluoroquinolone, Glycopeptide, MLS, Nitromidazole, Nucleoside, Peptide, Phenicol, Phosphonic acid, Rifamycin, Sulfonamide and Tetracycline. Additionally drug resistance against Biocides. | 16 | Detection in at least one environment amongst WWTP inflow, WWTP outflow, farm pond or freshwater lakes (water or sediments) | This study |
| 4 | **DeepARG** | AMRfinderplus DB | Aminoglycosides, Beta-lactams, Diaminopyrimidine, Fluoroquinolone, Glycopeptide, MLS, Multidrug, Nucleoside, Peptide, Phenicol, Phosphonic acid, Rifamycin, Sulfonamide and Tetracycline. Additionally drug resistance against Aminocoumarin. | 15 | Detection in at least one environment amongst WWTP inflow, WWTP outflow, farm pond or freshwater lakes (water or sediments) | This study |
| 5 | **Staramr** | ResFinder DB | Aminoglycosides, Beta-lactams, Diaminopyrimidine, Fluoroquinolone, MLS, Nitromidazole, Peptide, Phenicol, Phosphonic acid, Rifamycin, Sulfonamide and Tetracycline. | 12 | Detection in at least one environment amongst WWTP inflow, WWTP outflow, farm pond or freshwater lakes (water or sediments) | This study |
| 6 | **ResFinder v2.1 (web tool)** | not specified | Aminoglycosides, Beta-lactams, Cephalosporins, Folate Pathway Inhibitors (Sulfonamides and Diaminopyrimidines) and Tetracycline | 6 | Salmonella strains collected from Broiler Chickens | Cooper et al., 2020 |
| 7 | **KMA v1.17** | ResFinder, NCBI | Aminoglycosides, Beta-lactams, Cephalosporins, Folate Pathway Inhibitors (Sulfonamides and Diaminopyrimidines) and Tetracycline | 6 | Salmonella strains collected from Broiler Chickens | Cooper et al., 2021 |

| | | | | | | |
|---|---|---|---|---|---|---|
| 8 | **SRST2** | ResFinder, ARG-Annot, NCBI | Aminoglycosides, Beta-lactams, Cephalosporins, Penicillins and Tetracycline | 5 | Salmonella strains collected from Broiler Chickens | Cooper et al., 2022 |
| 9 | **RGI** | CARD | Aminoglycosides, Beta-lactams, Cephalosporins, Penicillins and Tetracycline | 5 | Salmonella strains collected from Broiler Chickens | Cooper et al., 2023 |
| 10 | **ABRICATE** | ResFinder DB, NCBI, ARG-ANNOT, CARD and MEGARes | Aminoglycosides, Beta-lactams, Fluoroquinolone, MLS, Efflux pump transporter (not an CARD drug class) and Tetracycline | 6 | Multiple sources (sewage, monkey, human, potable water and chicken) | Gomes et al., 2023 |
| 11 | **RGI** | CARD | Aminoglycosides, Beta-lactams, Fluoroquinolone, MLS, Efflux pump transporter (not an ARO class) and Tetracycline | 6 | Multiple sources (sewage, monkey, human, potable water and chicken) | Gomes et al., 2023 |
| 12 | **AMRPlusPlus** | MEGARes | Aminoglycosides*, Beta-lactams, Fluoroquinolone, MLS*, Phenicols*, Tetracycline and Diaminopyrimidines | 4 accurately; 3 low precission* | 500 isolates according to the following criteria: sequenced using Illumina platform; available AMR resistance profiles determined by phenotypic AST; available NCBI BioProject; and other parameters stated in the publication | Marini et al., 2022 |
| 13 | **Meta-MARC** | MEGARes | Aminoglycosides*, Beta-lactams, Fluoroquinolone*, MLS*, Phenicols*, Tetracycline and Diaminopyrimidines* | 2 accurately; 5 low precission* | 500 isolates according to the following criteria: sequenced using Illumina platform; available AMR resistance profiles determined by phenotypic AST; available NCBI BioProject; and other parameters stated in the publication | Marini et al., 2022 |
| 14 | **KARGA** | MEGARes, or any other fasta AMR DB | Aminoglycosides, Beta-lactams, Fluoroquinolone, MLS, Phenicols, Tetracycline and Diaminopyrimidines | 7 accurately | 500 isolates according to the following criteria: sequenced using Illumina platform; available AMR resistance profiles determined by phenotypic AST; available NCBI BioProject; and other parameters stated in the publication | Marini et al., 2022 |
| 15 | **DeepARG** | DeepARG-DB | Aminoglycosides*, Beta-lactams, Fluoroquinolone*, MLS, Phenicols* and Tetracycline. | 2 accurately; 5 low precission* | 500 isolates according to the following criteria: sequenced using Illumina platform; available AMR resistance profiles determined by phenotypic AST; available NCBI BioProject; and other parameters stated in the publication | Marini et al., 2022 |
| 16 | **ResFinder** | ResFinder DB , PointFinder DB | Aminoglycosides, Beta-lactams*, Fluoroquinolone, MLS, Phenicols, Tetracycline and Diaminopyrimidines | 6 accurately; 1 low precission* | 500 isolates according to the following criteria: sequenced using Illumina platform; available AMR resistance profiles determined by phenotypic AST; available NCBI BioProject; and other parameters stated in the publication | Marini et al., 2022 |

| | | | | | | |
|---|---|---|---|---|---|---|
| 17 | **Staramr** | ResFinder DB, PointFinder DB, PlasmidFinder DB | Aminoglycosides, Cephalosporin, Diaminopyrimidine, Phosphonic acid antibiotic, MLS, Nitrofuran, Penicillin, Phenicol, Fluoroquinolone, Sulfonamides, Tetracycline and Trimethoprim. | 12 | Simulated highly and low resistant mock community based on sequenced strains with known phenotypes. Following the requisites: (1) the strain had extensive antibiotic susceptibility CLSI or EUCAST testing data, (2) the strain was isolated from human tissue, (3) the strain was the cause of a clinical infection, (4) the FASTA was available to download from NCBI BioSample Database. | Wissel A et al., 2023 |
| 18 | **Srax** | CARD by default | Aminoglycosides, Beta-lactams, Diaminopyrimidine, MLS, Oxazolidinone, Fluoroquinolone. | 6 | Simulated highly and low resistant mock community based on sequenced strains with known phenotypes. Following the requisites: (1) the strain had extensive antibiotic susceptibility CLSI or EUCAST testing data, (2) the strain was isolated from human tissue, (3) the strain was the cause of a clinical infection, (4) the FASTA was available to download from NCBI BioSample Database. | Wissel A et al., 2024 |
| 19 | **Shortbread** | AMR gene marker database from 849 AR protein families from the ARDB19 and independent curation | Aminoglycosides, Beta-lactams, MLS, Fluoroquinolone and Tetracycline. | 5 | Simulated highly and low resistant mock community based on sequenced strains with known phenotypes. Following the requisites: (1) the strain had extensive antibiotic susceptibility CLSI or EUCAST testing data, (2) the strain was isolated from human tissue, (3) the strain was the cause of a clinical infection, (4) the FASTA was available to download from NCBI BioSample Database. | Wissel A et al., 2025 |
| 20 | **RGI** | CARD | Aminoglycosides, Beta-lactams, Glycopeptide, Cephalosporin, Diaminopyrimidine, Phosphonic acid antibiotic, Antibacterial free fatty acids, Glycopeptide, MLS, Nitrofuran, Nitroimidazole, Nucleoside, Oxazolidinone, Peptide, Phenicol, Pleuromutilin, Fluoroquinolone, Rhodamine (not an CARD drug class), Rifamycin, Sulfonamides and Tetracycline. Additionally drug resistance against Biocides. | 22 | Simulated highly and low resistant mock community based on sequenced strains with known phenotypes. Following the requisites: (1) the strain had extensive antibiotic susceptibility CLSI or EUCAST testing data, (2) the strain was isolated from human tissue, (3) the strain was the cause of a clinical infection, (4) the FASTA was available to download from NCBI BioSample Database. | Wissel A et al., 2026 |
| 21 | **Resfinder 4** | ResFinder 4 DB | Aminoglycosides, Beta-lactams, Glycopeptide, Diaminopyrimidine, Phosphonic acid antibiotic, MLS, Nitrofuran, Phenicol, Fluoroquinolone, Sulfonamides, Tetracycline and Trimethoprim. Additionally drug resistance against Biocides. | 13 | Simulated highly and low resistant mock community based on sequenced strains with known phenotypes. Following the requisites: (1) the strain had extensive antibiotic susceptibility CLSI or EUCAST testing data, (2) the strain was isolated from human tissue, (3) the strain was the cause of a clinical | Wissel A et al., 2027 |

| | | | | | | |
|---|---|---|---|---|---|---|
| | | | | | infection, (4) the FASTA was available to download from NCBI BioSample Database. | |
| 22 | **fARGene** | Hidden Markov models for quinolone, tetracycline, and beta lactamases | Beta-lactams, Fluoroquinolone, Tetracycline and Trimethoprim. | 4 | Simulated highly and low resistant mock community based on sequenced strains with known phenotypes. Following the requisites: (1) the strain had extensive antibiotic susceptibility CLSI or EUCAST testing data, (2) the strain was isolated from human tissue, (3) the strain was the cause of a clinical infection, (4) the FASTA was available to download from NCBI BioSample Database. | Wissel A et al., 2028 |
| 23 | **DeepARG** | DeepARG-DB | Aminoglycosides, Beta-lactams, Glycopeptide, Cephalosporin, Diaminopyrimidine, Phosphonic acid antibiotic, Glycopeptide, MLS, Nitroimidazole, Peptide, Phenicol, Fluoroquinolone, Sulfonamides, Tetracycline. Additionally drug resistance against Biocides. | 15 | Simulated highly and low resistant mock community based on sequenced strains with known phenotypes. Following the requisites: (1) the strain had extensive antibiotic susceptibility CLSI or EUCAST testing data, (2) the strain was isolated from human tissue, (3) the strain was the cause of a clinical infection, (4) the FASTA was available to download from NCBI BioSample Database. | Wissel A et al., 2029 |
| 24 | **AMRfinderplus** | AMRfinderplus DB | Aminoglycosides, Beta-lactams, Glycopeptide, Cephalosporin, Diaminopyrimidine, Phosphonic acid antibiotic, Antibacterial free fatty acids, MLS, Nitrofuran, Nucleoside, Peptide, Phenicol, Fluoroquinolone, Sulfonamides, Tetracycline and Trimethoprim. Additionally drug resistance against Biocides and Metals. | 18 | Simulated highly and low resistant mock community based on sequenced strains with known phenotypes. Following the requisites: (1) the strain had extensive antibiotic susceptibility CLSI or EUCAST testing data, (2) the strain was isolated from human tissue, (3) the strain was the cause of a clinical infection, (4) the FASTA was available to download from NCBI BioSample Database. | Wissel A et al., 2030 |
| 25 | **ABRICATE** | ResFinder DB, NCBI, ARG-ANNOT, CARD and MEGARes | Aminoglycosides, Beta-lactams, Cephalosporin, Diaminopyrimidine, Phosphonic acid antibiotic, Glycopeptide, MLS, Phenicol, Fluoroquinolone, Sulfonamides, Tetracycline and Trimethoprim. | 12 | Simulated highly and low resistant mock community based on sequenced strains with known phenotypes. Following the requisites: (1) the strain had extensive antibiotic susceptibility CLSI or EUCAST testing data, (2) the strain was isolated from human tissue, (3) the strain was the cause of a clinical infection, (4) the FASTA was available to download from NCBI BioSample Database. | Wissel A et al., 2031 |

**Table S3. Number of ARG hits detected for each intersection of environment (higher number of shared ARGs).** Open reading frames (ORFs) were annotated for antibiotic resistance genes (ARGs) and only those present in more than one environment were selected. Then the number of ARG hits for each intersection was listed in decreasing order. Intersecting environments with only 1 ARG are shown in Table S5.

| **Intersections between environments** | **Number of ARG hits** |
|---|---|
| WWTP inflow and outflow | 144 |
| WWTP inflow | 70 |
| WWTP inflow and outflow and urban water | 23 |
| Urban and rural sediments | 21 |
| WWTP inflow and outflow and urban sediment | 16 |
| WWTP inflow and outflow and farm pond | 7 |
| WWTP inflow and outflow, urban water and urban sediment | 6 |

| | |
|---|---|
| WWTP inflow and outflow, rural and urban sediment | 5 |
| WWTP inflow and outflow and rural sediment | 5 |
| WWTP inflow and outflow, farm pond and urban water | 5 |
| WWTP inflow and urban sediment | 2 |
| WWTP inflow and farm pond | 2 |
| Rural sediment | 2 |

**Table S4. Kruskal-Wallis statistics for those AMR classes which showed significant differences in the mean rank of the reads per kilobase per million mapped reads normalized by single copy marker genes (RPKM-SCMG) when compared between environments.** This table supports boxplots of Fig.3 (meaning a p-value adjusted > 0.05). Open reading frames (ORFs) were annotated for antibiotic resistance genes (ARGs) and the mean rank of the RPKM counts was computed by mapping the high-quality reads to the ORFs. This table also includes those AMR classes that show a non-adjusted p-value < 0.05. P-value adjusted significance {* < 0.05, ** < 0.01, *** < 0.001, **** < 0.0001}.

| ARG_class | .y. | n | statistic | df | p | method | p.adj | p.adj.signif |
|---|---|---|---|---|---|---|---|---|

| ARG_class | .y. | n | statistic | df | p | method | p.adj | p.adj.signif |
|---|---|---|---|---|---|---|---|---|
| aminoglycoside | count | 177 | 49.63 | 5 | 1.64e-09 | Kruskal-Wallis | 3.77e-08 | **** |
| beta-lactam | count | 137 | 34.99 | 4 | 4.66e-07 | Kruskal-Wallis | 1.07e-05 | **** |
| drug_biocide_resistance | count | 127 | 57.39 | 5 | 4.20e-11 | Kruskal-Wallis | 9.66e-10 | **** |
| mls | count | 81 | 24.58 | 4 | 6.09e-05 | Kruskal-Wallis | 1.40e-03 | ** |
| tetracycline | count | 52 | 33.54 | 5 | 2.93e-06 | Kruskal-Wallis | 6.73e-05 | **** |

**Table S5. Dun test statistics for the pairwise mean rank comparison of those AMR classes which showed significant differences in the mean rank of the reads per kilobase per million mapped reads normalized by single copy marker genes (RPKM-SCMG) when compared between environments.** This table supports boxplots of Fig.3 (meaning a p-value adjusted > 0.05). Open reading frames (ORFs) were annotated for antibiotic resistance genes (ARGs) and the mean rank of the RPKM counts was computed by mapping the high-quality reads to the ORFs. This table also includes those AMR classes that show a non-adjusted p-value < 0.05. P-value adjusted significance {* < 0.05, ** < 0.01, *** < 0.001, **** < 0.0001}.

| **ARG_class** | **.y.** | **group1** | **group2** | **n1** | **n2** | **statistic** | **p** | **p.adj** | **p.adj.signif** |
|---|---|---|---|---|---|---|---|---|---|
| aminoglycoside | count | WWTP-Inflow | Farm-Pond | 36 | 5 | -4.91 | 8.82e-07 | 5.73e-05 | **** |
| aminoglycoside | count | WWTP-Inflow | Urban-Sediment | 36 | 55 | -5.24 | 1.59e-06 | 1.03e-04 | **** |
| aminoglycoside | count | WWTP-Inflow | Urban-Water | 36 | 36 | -4.56 | 4.96e-06 | 3.22e-04 | *** |
| aminoglycoside | count | WWTP-Outflow | Farm-Pond | 30 | 5 | -3.42 | 6.24e-04 | 4.03e-02 | * |
| aminoglycoside | count | Farm-Pond | Rural-Sediment | 5 | 18 | 4.35 | 5.42e-04 | 3.52e-02 | * |

| ARG_class | .y. | group1 | group2 | n1 | n2 | statistic | p | p.adj | p.adj.signif |
|---|---|---|---|---|---|---|---|---|---|
| beta-lactam | count | WWTP-Inflow | Urban-Sediment | 54 | 19 | -4.72 | 2.35e-06 | 1.53e-04 | *** |
| beta-lactam | count | WWTP-Inflow | Rural-Sediment | 54 | 19 | -3.92 | 8.61e-05 | 5.60e-03 | ** |
| beta-lactam | count | WWTP-Outflow | Urban-Sediment | 49 | 19 | -4.02 | 5.69e-05 | 3.70e-03 | ** |
| beta-lactam | count | WWTP-Outflow | Rural-Sediment | 49 | 19 | -3.37 | 7.26e-04 | 4.72e-02 | * |
| drug_biocide_resistance | count | WWTP-Outflow | WWTP-Inflow | 61 | 61 | -4.36 | 1.67e-05 | 1.19e-03 | *** |
| drug_biocide_resistance | count | Farm-Pond | Rural-Sediment | 5 | 18 | -3.99 | 6.53e-02 | 4.24e-04 | ** |
| drug_biocide_resistance | count | Urban-Sediment | Rural-Sediment | 61 | 18 | -4.93 | 7.96e-06 | 5.17e-04 | **** |

| ARG_class | .y. | group1 | group2 | n1 | n2 | statistic | p | p.adj | p.adj.signif |
|---|---|---|---|---|---|---|---|---|---|
| drug_biocide_resistance | count | WWTP-Inflow | Urban-Water | 61 | 61 | -4.13 | 3.49e-04 | 2.26e-02 | ** |
| tetracycline | count | WWTP-Inflow | Farm-Pond | 36 | 5 | -3.55 | 3.83e-04 | 2.49e-02 | * |
| tetracycline | count | WWTP-Inflow | Urban-Sediment | 18 | 18 | -4.05 | 4.92e-06 | 3.19e-04 | ** |
| tetracycline | count | WWTP-Inflow | Rural-Sediment | 18 | 18 | -3.50 | 4.49e-04 | 2.91e-02 | * |

**Table S6. ARG intersection for those environments which share only one antibiotic resistance gene (ARG).** Open reading frames (ORFs) were annotated for antibiotic resistance genes (ARGs) and only those combinations of environments which shared 1 ARG were selected. Then the number of ARG hits for each intersection was listed.

| Intersections between environments | Number of ARG hits |
|---|---|
| Rural sediment, urban water and urban sediment | 1 |

| | |
|---|---|
| WWTP outflow | 1 |
| WWTP outflow and urban sediment | 1 |
| WWTP outflow and urban sediment and rural sediments | 1 |
| WWTP inflow and rural sediment | 1 |
| WWTP inflow, urban water and rural sediment | 1 |
| WWTP inflow and outflow, farm pond and rural sediment | 1 |
| WWTP inflow and outflow, rural sediment and urban water | 1 |
| WWTP inflow and outflow, urban sediment and farm pond | 1 |
| WWTP inflow and outflow, urban sediment, urban water, rural sediment | 1 |
| WWTP inflow and outflow, urban sediment, urban water, rural sediment and farm pond | 1 |

**Table S7. AMR class abundance comparison between the WWTP Inflow and the WWTP Outflow.** Open reading frames (ORFs) were annotated for antibiotic resistance genes (ARGs) and only those belonging to the WWTP inflow and WWTP outflow and, which had an AMR class assigned, were listed and sorted by abundance (RPKM-SCMG counts).

| | group | ARG_class | counts | | group | ARG_class | counts |
|---|---|---|---|---|---|---|---|
| 1 | WWTP-Inflow | sulfonamide | 62.11926176 | 1 | WWTP-Outflow | sulfonamide | 51.46090567 |
| 2 | WWTP-Inflow | tetracycline | 43.29441272 | 2 | WWTP-Outflow | MLS | 9.313243386 |
| 3 | WWTP-Inflow | MLS | 18.6230147 | 3 | WWTP-Outflow | beta-lactam | 8.913506546 |
| 4 | WWTP-Inflow | cephalosporin | 17.02206724 | 4 | WWTP-Outflow | tetracycline | 8.806977195 |
| 5 | WWTP-Inflow | aminoglycoside | 13.60019685 | 5 | WWTP-Outflow | aminoglycoside | 5.835370769 |
| 6 | WWTP-Inflow | beta-lactam | 11.10864217 | 6 | WWTP-Outflow | diaminopyrimidine | 3.417023701 |
| 7 | WWTP-Inflow | phenicol | 10.97504052 | 7 | WWTP-Outflow | multidrug | 3.065306204 |
| 8 | WWTP-Inflow | fluoroquinolone | 9.853703689 | 8 | WWTP-Outflow | fluoroquinolone | 2.219676743 |
| 9 | WWTP-Inflow | diaminopyrimidine | 7.6070189 | 9 | WWTP-Outflow | cephalosporin | 2.196473552 |
| 10 | WWTP-Inflow | peptide | 3.442193257 | 10 | WWTP-Outflow | phenicol | 2.028349205 |
| 11 | WWTP-Inflow | multidrug | 3.248614903 | 11 | WWTP-Outflow | peptide | 1.55388725 |
| 12 | WWTP-Inflow | biocide resistance | 3.220638809 | 12 | WWTP-Outflow | biocide resistance | 1.14984574 |
| 13 | WWTP-Inflow | rifamycin | 2.865307999 | 13 | WWTP-Outflow | phosphonic acid | 1.14088951 |
| 14 | WWTP-Inflow | phosphonic acid | 2.421449837 | 14 | WWTP-Outflow | rifamycin | 1.082781457 |
| 15 | WWTP-Inflow | nitroimidazole | 2.248053219 | 15 | WWTP-Outflow | biocide and metal resistance | 0.747727273 |
| 16 | WWTP-Inflow | biocide and metal resistance | 1.406362499 | 16 | WWTP-Outflow | nucleoside | 0.622857143 |

| | | | |
|---|---|---|---|
| 17 | WWTP-Inflow | glycopeptide | 1.141990919 |
| 18 | WWTP-Inflow | nucleoside | 0.659293726 |
